# A Rare Variant in *TRIOBP* Linked to Occupational Noise Exposure in Meniere Disease

**DOI:** 10.1101/2025.07.23.25332035

**Authors:** Pablo Cruz-Granados, Giselle Bianco-Bortoletto, Yuzhong Zhang, Prathamesh T. Nadar-Ponniah, Kiana Bagheri-Loftabad, Edi Lúcia Sartorato, Ines Sanchez-Sellero, Andres Soto-Varela, Jose A. Lopez-Escamez

**Affiliations:** Meniere Disease Neuroscience Research Program, Faculty of Medicine & Health, School of Medical Sciences, The Kolling Institute, University of Sydney, Sydney, New South Wales, Australia; Laboratory of Human Molecular Genetics, Center for Molecular Biology and Genetic Engineering - CBMEG, Programa de Pós Graduação em Ciências Médicas, Faculty of Medical Sciences, State University of Campinas - UNICAMP, Sao Paulo, Brazil; Department of Otorhinolaryngology-Head & Neck Surgery, West China Hospital of Sichuan University, Chengdu, China; Division of Toxicology, Department of Forensic Sciences, Pathology, Gynecology and Obstetrics, and Pediatrics, School of Medicine, Forensic Sciences Institute, Universidade de Santiago de Compostela, 15782 Santiago de Compostela, Spain; Division of Neurotology, Department of Otorhinolaryngology, Complexo Hospitalario Universitario de Santiago de Compostela, 15706 Santiago de Compostela, Spain; Department of Surgery and Medical-Surgical Specialities, School of Medicine, Universidade de Santiago de Compostela, 15782 Santiago de Compostela, Spain; Health Research Institute of Santiago (IDIS), 15706 Santiago de Compostela, Spain; Otology & Neurotology Group CTS495, Division of Otolaryngology, Department of Surgery, Instituto de Investigación Biosanitaria, ibs.GRANADA, Universidad de Granada, Granada, Spain; Sensorineural Pathology Programme, Centro de Investigación Biomédica en Red en Enfermedades Raras, CIBERER, Madrid, Spain; Ear Science Institute Australia, Nedlands, Western Australia, Australia

**Author notes:** Correspondence: Prof. Jose A. Lopez-Escamez Faculty of Medicine & Health, School of Medical Sciences, University of Sydney The Kolling Institute 10 Westbourne St, St Leonards, NSW 2064 Sydney Australia.

**Keywords:** Genetics, Hearing Loss, Inner Ear, Meniere Disease, Noise

## Abstract

Meniere disease (MD) is an inner ear disorder characterised by episodic vertigo, tinnitus, and sensorineural hearing loss. Previous sequencing studies have identified rare mutations in 16 genes expressed in hair cells, many of which are involved in maintaining the structure and stability of stereocilia. Experimental evidence supports that noise-induced hearing loss is associated with structural damage in the hair cell stereocilia. We hypothesise that rare variants in genes expressed in hair cell stereocilia may predispose individuals to MD with occupational noise exposure. To explore this, we analysed whole exome sequencing data from MD patients with documented histories of occupational noise exposure. A rare missense variant chr22:37769343C>T in the *TRIOBP* gene was identified in three individuals (OR=1,846 (203-8192), p=3.39x10^-5^). In silico protein modelling suggests this variant (p.R2273C) interacts with the Actin C chain, potentially affecting cytoskeletal integrity in hair cells. Immunolabelling confirmed *TRIOBP* expression in the stereocilia rootlets of cochlear and vestibular hair cells, supporting its structural function in both sensory epithelia.

Furthermore, retrieved MD single-cell ATAC-seq data revealed reduced chromatin accessibility at the *RFX3* transcription factor locus, suggesting a transcriptional *TRIOBP* downregulation. Our results support that occupational noise exposure may trigger MD in the carriers of *TRIOBP* mutations.

## Introduction

Menière’s Disease (MD) is a lifelong disease characterised by episodes of vertigo, sensorineural hearing loss (SNHL) and tinnitus.^1^ The condition is associated with the accumulation of endolymph in the cochlear duct, termed endolymphatic hydrops (EH), associated with SNHL. Clinical research showing familial clustering^2^ and exome sequencing studies in multi-case families have provided evidence to associate 15 genes with MD,^3^ including autosomal dominant (*DTNA*, *TECTA*, *GJD3*) ^2,4,5^ and compound recessive inheritance (*OTOG*)^6^ Moreover, genomic data from different studies have shown that sporadic (non-familial) MD patients share around 27% of the variants and 28% of genes with familial MD.^7,8^ Most of the genes linked to MD encode large structural proteins in the organ of Corti, either in the tectorial membrane or the hair cell stereocilia, including *MYO7A* or *TECTA* or *OTOG* previously associated with non-syndromic SNHL.^9–11^ Knock-out models of these genes have confirmed structural abnormalities in the tectorial membrane or the hair cell stereocilia bundle.^12^

Noise-induced hearing loss (NIHL), including blast injury has been associated with EH in animal models,^13^ supporting the hypothesis that noise trauma could lead to histological damage with acute inflammatory changes in the cochlear structures such as the stria vascularis and the hair cell stereocilia in the organ of Corti.^14^ Moreover, loud sound could deplete Ca^2+^ from tectorial membrane leading to nonfunctional mechanotransduction channels and loss of hearing sensitivity.^15^

The *TRIOBP* gene encodes for TRIO and F-actin-binding protein, a structural protein is found in several organs across the body, with isoforms 4 and 5 found primarily at the base of the hair cell stereocilia.^16–18^ Mutations in *TRIOBP* are associated with nonsyndromic recessive SNHL (DFNB28) across different populations.^17,19,20^ The *TRIOBP*-encoded protein isoform 6 is the longest isoform found in humans, but its function is not well known since *TRIOBP*-6 is not expressed in murine species.^21^

We hypothesise that hippomorphic mutations in structural proteins of the hair cell stereocilia changing the protein stability could reduce the tolerance to endolymphatic pressure changes leading to a loss of protein interactions and structural changes involving mechanotransduction. To explore this hypothesis, we retrieved exome sequencing data from a cohort of MD individuals with and without a history of occupational noise exposure to search for rare variants associated with noise-exposure in MD.

## Material and Methods Human and Animal Ethics

The Human Ethics Research Committee (2023/HE000199) from The University of Sydney approved the protocol for this study. A written inform consent was obtained from all participants to donate blood samples, extract DNA for its sequencing and perform genetic analyses. This work was performed under the standards of the Declaration of Helsinki.

The Animal Ethics Research Committee (approval no. 2023/2388) from The University of Sydney and the Animal Ethical and Welfare (approval no. 20240219057) from West China Hospital of Sichuan University evaluated and approved the protocol for this study.

## Patient Selection

Clinical data pertaining occupational noise exposure across a three-year study was retrieved from 77 individuals from Spain with diagnosed MD.^22^ Participants were categorised as exposed to noise and/or vibrations (n = 39) or not exposed to noise and/or vibrations (n = 38). These risks were listed for their occupation (International Standard Classification of Occupations, 2008 (ISCO-08) in Spain’s Social Security National Institute (SSNI) guide.

We obtained whole exome sequencing (WES) data from 17 MD individuals that were categorised as exposed to occupational noise and/or vibrations, and 17 MD categorised as not exposed.

## Genomic Datasets

Previously published exome and genome sequencing data from MD participants were retrieved.^7,8^ Gene burden analyses (GBA) were performed to generate a list of MD genes with excess of rare variants overlapping in these datasets.

Hearing loss associated genes were retrieved from the Deafness Variation Database.^23^ In addition, we also retrieved single-cell RNA sequencing differentially expressed genes (DEG) found in the stria vascularis (SV), marginal cells (MC), intimidate cells (IMC) and basal cells (BC) of P30 mice.^24^

Finally, a list of genes from WES data associated with noise-induce hearing loss was retrieved and used to generate a list of NIHL genes for further analyses.^25^

## Identification of Rare Variants in the Occupational Noise Cohort

Single nucleotide variants and short indels in the noise and non-noise cohorts were retained if their allelic frequency (AF) <0.05 in Non-Finnish European (NFE) from gnomAD and found at least in two out of the 17 (11.7%) individuals in their respective category.

We retrieved variants from the noise cohort that overlapped with the MD list, the DVD list, and the SV list; NIHL gene list was used as an internal control. The same variant extraction was done for the MD cohort without noise-exposure. The retrieved gene variants from the noise and non-noise exposed cohort were compared, and replicated variants were discarded.

Long non-coding RNA (lncRNA) variants were also extracted from individuals with rare variants in the noise cohort. LncRNAs that regulate the expression of candidate genes, as well as those that modulate the activity of co-expressed genes, were retained for further analyses.

## Statistical Analysis

Allelic frequencies were obtained for the three variants sets and were compared to the AFs of the NFE reference dataset from gnomAD v4.1.0^26^ to calculate the odds ratio (OR), 95% confidence interval (CI) and *p*-value adjusted using the Bonferroni correction.

## Chromatin accessibility and Methylation Profiling of Candidate Genes

Epigenetic information from MD patients’ peripheral blood mononuclear cells generated using Whole Genome Bisulphite Sequencing (WGBS)^27^ and pseudo-bulk ATAC sequencing (ATACseq)^28^ was retrieved form previously published studies. The WGBS dataset was used to determine differentially methylated regions, whereas the ATACseq dataset was used to determine chromatin accessibility regions. The ATACseq data was generated differential peak analysis of MD cases against a set of internal controls (HC) and external controls (EC).

## Pathogenicity and Splice Site Prediction

Splice sites were predicted using *SpliceAI*^29^ and *Pangolin*^30^ for selected proteins using the canonical and relevant sequences. An upstream and downstream window consisting of 500-base pair (bp) from the variant position was used. In addition, Exonic Splicing Enhancer (ESE), Exonic Splicing Silencers (ESS), Intronic Splicing Enhancers (ISE) and Intronic Splicing Silencers (ISS) were predicted using *Human Splice Finder Pro* (HSF Pro).^31^

Pathogenicity scores were predicted using CADD score, PolyPhen2^32^ and REVEL,^33^ and pathogenicity heatmaps were generated for missense variants in canonical sequences from AlphaMissese^34,35^ pathogenicity predictions. GnomAD v4.1.0 *Z*-scores were obtained to evaluate constraint regions around the variant of interest. The Variant Interpretation Platform for Genetic Hearing Loss^36^ was used to classify the pathogenicity of the variants using the American College of Medical Genetics and Genomics (ACMG) criteria adjusted to Genetic Hearing Loss.^37^

Constraint regions with an overload of missense variants in genes of interest were identified by calculating the density of variants in the coding sequence (CDS). A sliding window consisting of 100 bp upstream and 100 bp downstream for the chosen mutations was created. The missense variants were retrieved from gnomAD v.2.1 database^16^ for each population. Estimation of the high-density threshold was calculated based on the expected number of missense variants for each population defined in gnomAD v.2.1. Regions were classified as low-density if the computed density of the window was below the predicted density for each population.

## Protein Modelling

Wild type (wt) sequences of interest were retrieved from UniProt^38^ and PDB files were obtained from AlphaFold Protein Structure Database^35^ and OtoProtein2 Database.^39^ Atomic interactions of the wild type models were examined using ERRAT^40^ from Saves v6.1 server (https://saves.mbi.ucla.edu/). For proteins without experimentally resolved structures or models with a ERRAT score < 45, models were obtained using AlphaFold 3^41^ (*ab initio*) and Swiss-Model^42^ (by homology).

Mutated proteins were generated by homology from wild type templates using MODELLER v10.6^43^ and Swiss-Model. MODELLER bult-in quality controls (DOPE and GA341) were used to determine suitability of the model. The chosen model had the lowest DOPE score and a GA341 score close to one. Swiss-Model predicts protein 3D structures by homology modelling by aligning target sequences with reference templates from their database. The selection of the optimal model is based on QMEAN scoring, which is a combination of geometry, solvation and agreement with expected properties. The reliability of the models is further validated through the implementation of GMQE (Global Model Quality Estimate) and local *Z*-scores, which identify and highlight any unreliable regions.

External quality checks were performed on mutated proteins using ERRAT, WHATCHECK (https://swift.cmbi.umcn.nl/gv/whatcheck/index.html) and PROCHECK^44^ validation algorithms from Saves v6.1 server. WHATCHECK and PROCHECK were employed for assessing stereochemical quality of the models.

The chosen wt and mutated protein models were analysed for structural changes using PyMOL (*Schrodinger*, LLC. 2010. The PyMOL Molecular Graphics System, Version 3.0.). Finally, DynaMut2^45^ and MUPro^46^ were used for predicting protein stability on the mutant proteins.

## Protein-Protein Interactions

Prediction of functional partners was performed using STRING v12^47^ on the selected proteins, just using from “curated databases” and “experimentally determined” options. BioGRID v4.4 (https://wiki.thebiogrid.org/, accessed on the 10^th^ of February 2025) and HINT^48^ (https://hint.yulab.org/, accessed on the 27^th^ of February 2025) were used to determined experimentally protein-protein interactions for selected proteins. The gEAR portal^49^ *Human inner ear organoids - scRNAseq and snRNAseq dataset*^50^ was used to determine expression in inner ear of predicted interactions with selected proteins. Proteins that expressed a fold change >1 in hair cells were retained for further analyses.

The International Mouse Phenotyping Consortium (IMPC)^51,52^ was used to determine if gene knockout of functional patterns, mice developed hearing loss or vestibular dysfunction.

## Molecular Docking

Prediction of interaction sites between the proteins of interest and functional partners was performed using DockNet.^53^ ClusPro^54^ was used to assess the molecular docking of selected PDB structures. The quality of the model is assessed via consensus clustering (reproducibility across scoring functions) and interface-specific metrics such as pairwise residue contacts (e.g., CAPRI criteria). The selection of the optimal model was based on cluster population (highest-density centres) and balanced scoring (electrostatics + van der Waals + decoy discrimination).^55^ DynaMut2 was used to evaluate protein stability in the mutant proteins of interest once docked to functional partners.

## Histology and Immunolocalization

Mouse inner ear samples were prepared for whole-mount immunofluorescence as follows. After deep anaesthesia and decapitation, the inner ears were dissected and fixed in 4% paraformaldehyde (PFA) (Cat#: BL539A, Biosharp) at 4°C overnight.

The cochlear basilar membrane and vestibular macula were carefully dissected under a stereomicroscope, fixed for 30 minutes, and then decalcified in 10% EDTA for an additional 30 minutes. Tissues were permeabilized and blocked in a solution containing 1% Triton X-100 (Cat#: 1139ML100, BioFroxx) and 5% bovine serum albumin (BSA) (Cat#: SRE0098-10G, Sigma).

Samples were then incubated overnight at 4°C with rabbit anti-ninein primary antibody (1:500; Cat#: 13007-1-AP, Proteintech) or *TRIOBP* antibody (1:500, proteintech 161241-AP) diluted in a solution of 0.1% Triton X-100 and 5% BSA. The following day, samples were washed three times with 1× PBS containing 0.1% Triton X-100, then incubated for 2 hours at room temperature with secondary antibody (Donkey anti-Rabbit IgG; Cat#: A32795, Invitrogen), Goat anti rabbit Alexa Fluor™ 594 (Cat # ab150080) and Alexa Fluor™ 555 Phalloidin (Cat#: A34055, Invitrogen), Phalloidin-iFluor 488 (Cat # ab176753). After additional washing with 1× PBS containing 0.1% Triton X-100, nuclei were counterstained with DAPI (Cat#: C1006, Beyotime). Finally, samples were mounted either using anti-fade fluorescence mounting medium (Cat#: AB104135, Abcam) or Prolong Gold antifade mounting medium ( Cat #P36935) and imaged using a Leica Stellaris 5 confocal microscope.

## Results

### An Ultrarare Variant in the *TRIOBP* Gene is Associated to Occupational Noise Exposure in MD

A total of five rare missense variants across four different genes (*TRIOBP*, *CENPJ*, *PLEKHA7* and *SLC41A3*) were found to be significantly associated in the noise exposed MD cohort (Table 1). None of them were found in the MD cohort without history of occupational noise exposure. Two of these variants in the *CENPJ* gene (chr13:24912739T>C, p.H96A; chr13:24879282A>G), and one variant in the *PLEKHA7* gene (chr11:16789275G>A, p.R1060C) were also replicated in the MD exome dataset in four, five and eight individuals respectively. Interestingly, the ultrarare missense variant in the *TRIOBP* gene (chr22:37769343C>T, p.R2273C, AF=3.22x10^-5^) that was found in 2/17 (11%) MD individuals in the noise exposed cohort (OR=1846.76 [203.82-8192], *p*-value adjusted [*p*-val adj] = 3.39x10^-5^), was previously reported in the DVD. Finally, the 3’ UTR variant found the *SLC41A3* gene (chr3:126006491T>C) was found to be differentially expressed in the marginal cells of the SV.

**Table 1.** Rare variants in *TRIOBP*, *CENPJ*, *PLEKHA7* and *SLC41A3* genes found in the occupational noise exposure cohort.

| MD Noise Cohort |  |  |  |  |  |
| --- | --- | --- | --- | --- | --- |
| Gene | *TRIOBP* | *CENPJ* | *CENPJ* | *PLEKHA7* | *SLC41A3* |
| **Variant ID** | chr22:37769343C>T<br>rs747351398 | chr13:24912739T>C<br>rs61739263 | chr13:24879282A>G<br>rs2275939 | chr11:16789275G>A<br>rs143911467 | chr3:126006491T>C<br>rs9833685 |
| **Consequence** | missense_variant | missense_variant | downstream_gene_variant | missense_variant | 3_prime_UTR_variant |
| **Codons** | Cgc/Tgc | cAt/cGt | - | Cgc/Tgc | - |
| **Amino Acid Change** | p.Arg2273Cys | p.His96Arg | - | p.Arg1060Cys | - |
| **gnomADg_AF_nfe** | 3.22E<sup>-05</sup> | 0.000283 | 0.01 | 0.000661 | 0.00147 |
| **CSVS_AF** | 0 | 0.002 | 0.003 | 0.001 | 0.011 |
| **AF** | 0.0588 | 0.0588 | 0.0588 | 0.0588 | 0.0588 |
| **OR (CI 95%),** | 1,847 (204-8192), | 208 (24-829), | 80 (9.1-313), | 89 (0.33-350), | 40 (4.7-157), |
| **p-value** | 3.39x10<sup>-05</sup> | 0.00202 | 0.0136 | 0.0002 | 0.0065 |
| **n_het** | 2 | 2 | 2 | 2 | 2 |
| **n_hom** | 0 | 0 | 0 | 0 | 0 |
| **CADD_PHRED** | 26.7 | 0.18 | 10.01 | 29.9 | 0.2 |
| **HL ACMG classification** | Uncertain Significance | Benign | Benign | Uncertain Significance | Benign |

\* CSVS: Collaborative Spanish Variant Server
\*\* HL ACMG: Hearing Loss American College of Medical Genetics

Furthermore, rare variants in the genes *PRRC2C* (chr1:171536182A>G, p.M733V) and *NEURL4* (chr17:7321200T>A, p.E1091V) were found associated exclusively in the noise exposure MD cohort. These genes were previously associated with NIHL but had not been previously associated with MD (Table 2).

**Table 2.** Rare variants in *PPRC2C* and *NEURL4* genes found in the occupational noise exposure cohort.

| MD Noise Cohort Associated to NIHL |  |  |
| --- | --- | --- |
| Gene | *PPRC2C* | *NEURL4* |
| **Variant ID** | chr1:171536182A>G<br>rs138220849 | chr17:7321200T>A<br>rs145900596 |
| **Consequence** | missense_variant | missense_variant |
| **Codons** | Atg/Gtg | gAg/gTg |
| **Amino Acid Change** | p.Met733Val | p.Glu1091Val |
| **gnomADg_AF_nfe** | 4.11E<sup>-04</sup> | 1.76E<sup>-03</sup> |
| **CSVS_AF** | 0.003 | 0.012 |
| **AF** | 0.0588 | 0.0588 |
| **OR (CI 95%)** | 143 (16.6-557), | 33 (3.9-131), |
| **p-value** | 0.0025 | 0.05 |
| **n_het** | 1 | 1 |
| **n_hom** | 0 | 0 |
| **CADD_PHRED** | 23 | 24.5 |
| **HL ACMG classification** | Uncertain Significance | Benign |

Notably, the variant chr1:236524482A>G in the *LGALS8-AS1* lncRNA variant was found in the same two individuals with the *TRIOBP* variant and it was not observed in the MD non-noise cohort.

However, this variant with an AF = 0.04 in gnomAD-NFE, was not significantly associated (OR=1.35 [0.16-5.28], *p*-value adjusted >0.05).

## Identification of Epigenomic Modifications in *TRIOBP* in MD NIHL Cohort

Six differentially methylated CpGs (DMC) were found in the WGBS data and four differentially accessible (DA) peaks: one in the 3’ untranslated region (UTR), two in coding sequences in the pseudo-bulk ATACseq MD-HC data and one in the coding sequence for MD-EC data for *TRIOBP* (Table A.1). The DMC were between 39,646 and 57,778 base pairs upstream from our variant of interest. For the ATACseq there was only one range that was a significant 3’ UTR chr22-37696386- 37697412 (Fold Change [log2FC] = -1.50; *p*-val adj = 2.97x10^-04^).

Furthermore, DA peak data for *RFX3* transcription factor was retrieved as it regulated *TRIOBP* expression in inner ear hair cells.^56^ On the MD-HC data we found a GAP and 3’UTR regions, whereas on the MD-EC data we found a UTR regions. Both UTR regions were significantly downregulated in MD patients (log2FC = -1.22; *p*-val adj = 6.53x10^-24^ and log2FC = -1.07; *p*-val adj =1.19x10^-13^ respectively) (Table A.2).

### *TRIOBP* Carriers Clinical Phenotype

Patient 1 was in the seven-decade of life with a familial history of MD (Fig. 1A). They developed an episodic vestibular syndrome associated with right ear SNHL involving all frequencies in their late forties, without any additional comorbidity (autoimmune diseases or headache) (Fig. 1B). Their brother was in their early-sixties. They developed bilateral MD and has reported a history of migraine and Tumarkin otolithic syndrome.

**Figure 1.**
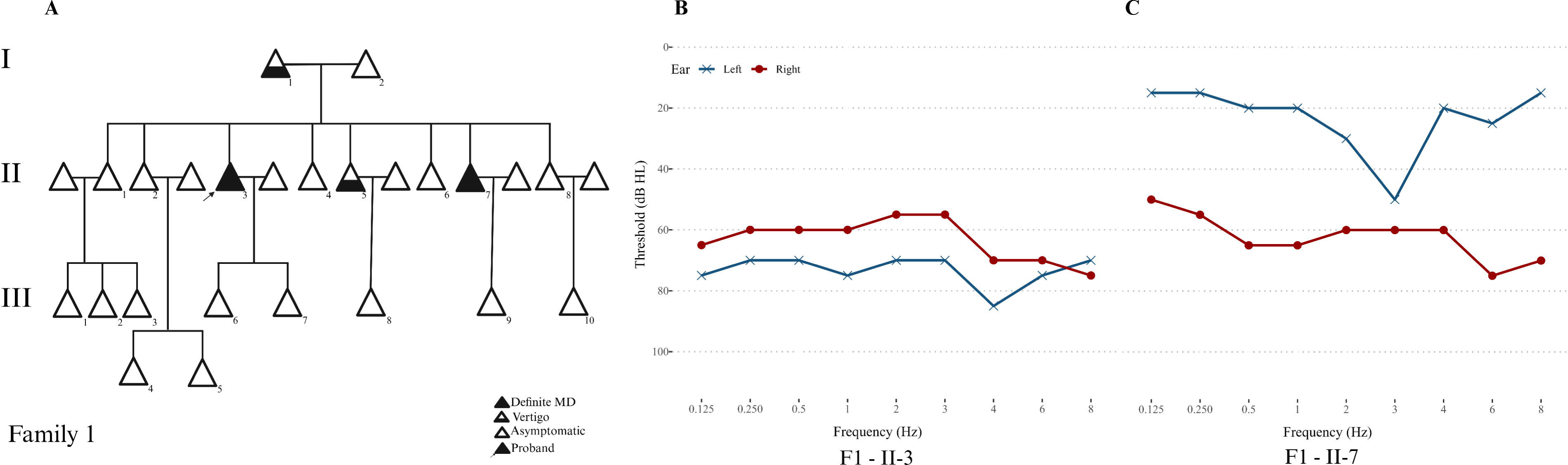
A) Pedigree of Family 1 with *TRIOBP* chr22:37769343C>T mutation. B) Proband II-3 air conducted hearing threshold audiogram with bilateral SNHL. C) Family member II-7 air conducted hearing threshold audiogram with unilateral SNHL in right ear. Left ear represented in blue; right in red.

The second patient was in the mid-eighties with a long history of episodic vertigo and unilateral SNHL in the left ear, involving also all frequencies (Figure A.1). This individual has no familial history, neither other relevant condition associated with MD.

## Pathogenicity Assessment

The *TRIOBP* chr22:37769343C>T variant was classified as Variant of Uncertain Significance according to the Hearing Loss-specific ACMG criteria.

No splice sites were detected for *TRIOBP* chr22:37769343C>T variant using *Pangolin* or *SpliceAI*. On the other hand, *HSF Pro* found that a new Donor Splice Site was generated around chr22:37769343C>T variant (**Table A.3**). The splice site starts a position 37769339 and finishes at position 37769347 in exon 22. Furthermore, five ESE sites were broken, three surrounding the variant and two at the variants’ position. Finaly two new ESS sites were generated in the periphery of the variant. The splice site, the ESE and the ESS were associated to the *TRIOBP* canonical transcript (ENST00000644935.1) and alternative transcript ENST00000403663.6 (**Table A.4**).

The distribution of missense variants along the *TRIOBP* CDS allowed us to define high-density regions (HDR) and low-density regions (LDR). We observed the distribution of these regions was similar across different populations (**Fig. 2A**). Interestingly, we found that *TRIOBP* chr22:37769343C>T (CDS position 6817) variant is found in a LDR region in the NFE population, but in HDR regions for South Asian (SAS), East Asian (EAS), Admixed American (AMR) and Finnish European (FIN) populations.

**Figure 2.**
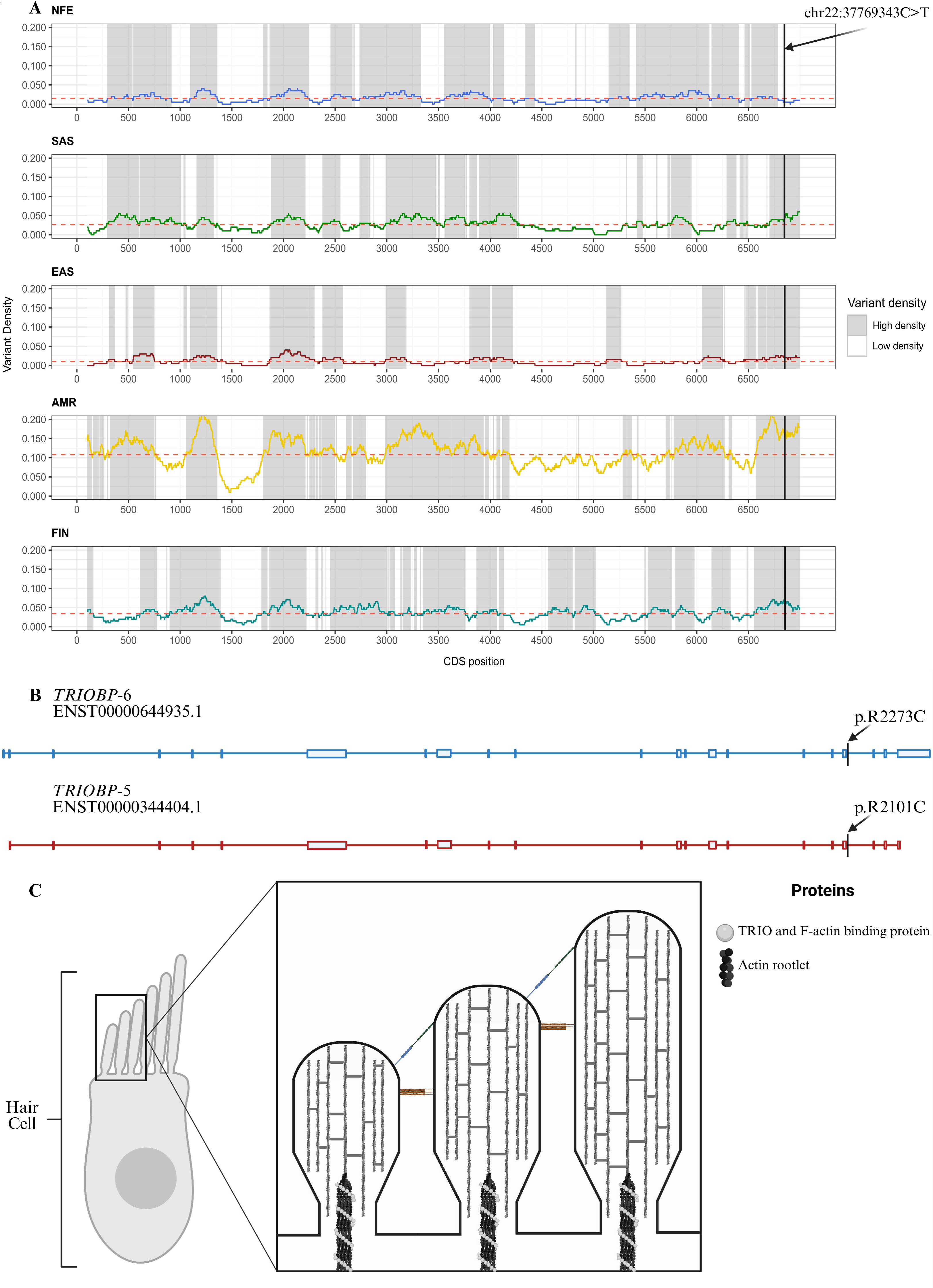
A) Low-density and high-density areas in *TRIOBP* CDS across different populations. B) *TRIOBP*-6 ENST00000644935 and *TRIOBP*-5 ENST00000344404 transcripts. C) *TRIOBP*-5 linking F-actin proteins in the stereocilia rootlets to add rigidity to the structure. .

### *TRIOBP* Protein Modelling

Modelling of wt TRIO and F-actin-binding protein (*TRIOBP*) was performed by homology using Swiss-Model, on *TRIOBP*-6 (canonical transcript, ENST00000644935.1) and *TRIOBP*-5 (ENST00000344404.10 transcript) isoforms as the variant is present in p.R2273C and p.R2101C respectively (**Fig. 2B**).

Modelling of both protein isoforms revealed the mutation is in a small loop region joining two alpha helices. AlphaMissense pathogenicity heatmap for the canonical sequence showed that the mutation p.R2273C is found in a benign site (**Figure A.2)**.

DynaMut2 predicted that the replacement of arginine for cysteine would stabilise both proteins. For *TRIOBP*-6 a change 0.22 kcalmol^-1^ was predicted, whereas a change of 0.17 kcalmol^-1^ was reported for *TRIOBP*-5. The replacement of an arginine for a cysteine at p.2101 in *TRIOBP*-5 added one more polar contact between p.C2101 and p.E2104, and lost its Van der Waals (VDW) interaction with p.C2099 while creating a hydrogen bond instead (**Fig. 3A**).

**Figure 3.**
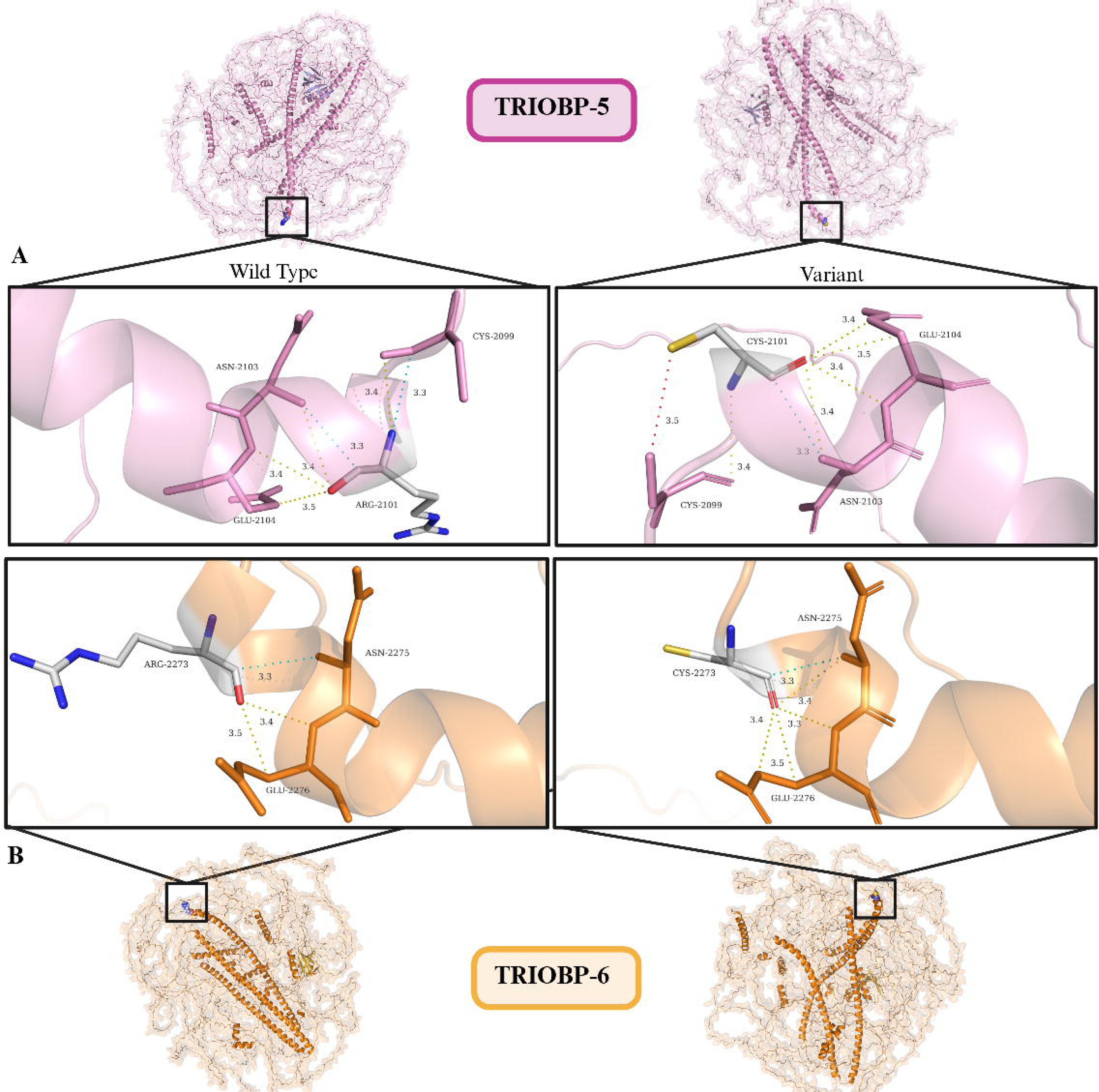
A) Wild type (left) and mutant (right) *TRIOBP*-5 protein model, zooming in at the atomic interactions of *TRIOBP*-5 p.2101. . B) Wild type (left) and mutant (right) *TRIOBP*-6 protein model, zooming in at the atomic interactions of *TRIOBP*-6 p.2273.

In contrast to *TRIOBP*-5, mutated *TRIOBP*-6 residue p.C2273 retained the wt residue interactions whereas additionally forming two new polar contacts, one with p.N2275 and another with p.E2276.

## *TRIOBP* Molecular Docking with Functional Partners

STRING predicted *TRIOBP* interactions with five proteins encoded by *PACSIN2*, *XIRP2*, *SPTAN1, SPTBN1*, and *PJVK*. On the other hand, BioGRID has reported 131 known protein-protein interaction with *TRIOBP*. Out of the 136 interactions, only four proteins were significantly expressed in hair cell organoids (*GSK3B*, *PCM1*, *SPTBN1* and *NIN*). Furthermore, IMPC *NIN*-knockout data shows that mice develop high frequency hearing loss.^51^

Docknet predicted low binding affinity between p.R2273 and p.R2101 in *wtTRIOBP-6* and wt*TRIOBP*-5 with *NIN*, *PCM1*, *GSK3B* or *SPTBN1* compared to other positions in the proteins (**Figures A.3 – A.10**). Ninein protein model (*NIN* gene) was generated by homology using Modeller and was able to dock to *TRIOBP*-6 but not *TRIOBP*-5 (**Figures A.11 – A.12**).

Given *TRIOBP*-6 lack of studies, and *TRIOBP*-5 known actin-binding properties, we investigated their interactions with the β-actin and filamentous (F-actin) forms (**Fig. 2C**). Protein models (**Figures A13–A18**) showed that wt and mutant *TRIOBP*-6 docked with both β-actin and F-actin in a different position to p.2273, similarly wt and mutant *TRIOBP*-5 docked at a different site on F-actin. However structural analysis utilising the PyMOL software programme identified two polar interactions between *TRIOBP*-5 residue p.2101 and β-actin p.363 (**Fig. 4**). In the monomer structure, the p.R2101C mutation shortened the distance of one of these contacts. Furthermore, computational stability analysis predicted a delta change (ΔΔG_pred) of 0.72 kcal^-1^ for this mutation. Docknet predicted a probability of interaction between *TRIOBP*-5 and *ACTB* was of 0.53 (**Figures A.19 – A.20**).

**Figure 4.**
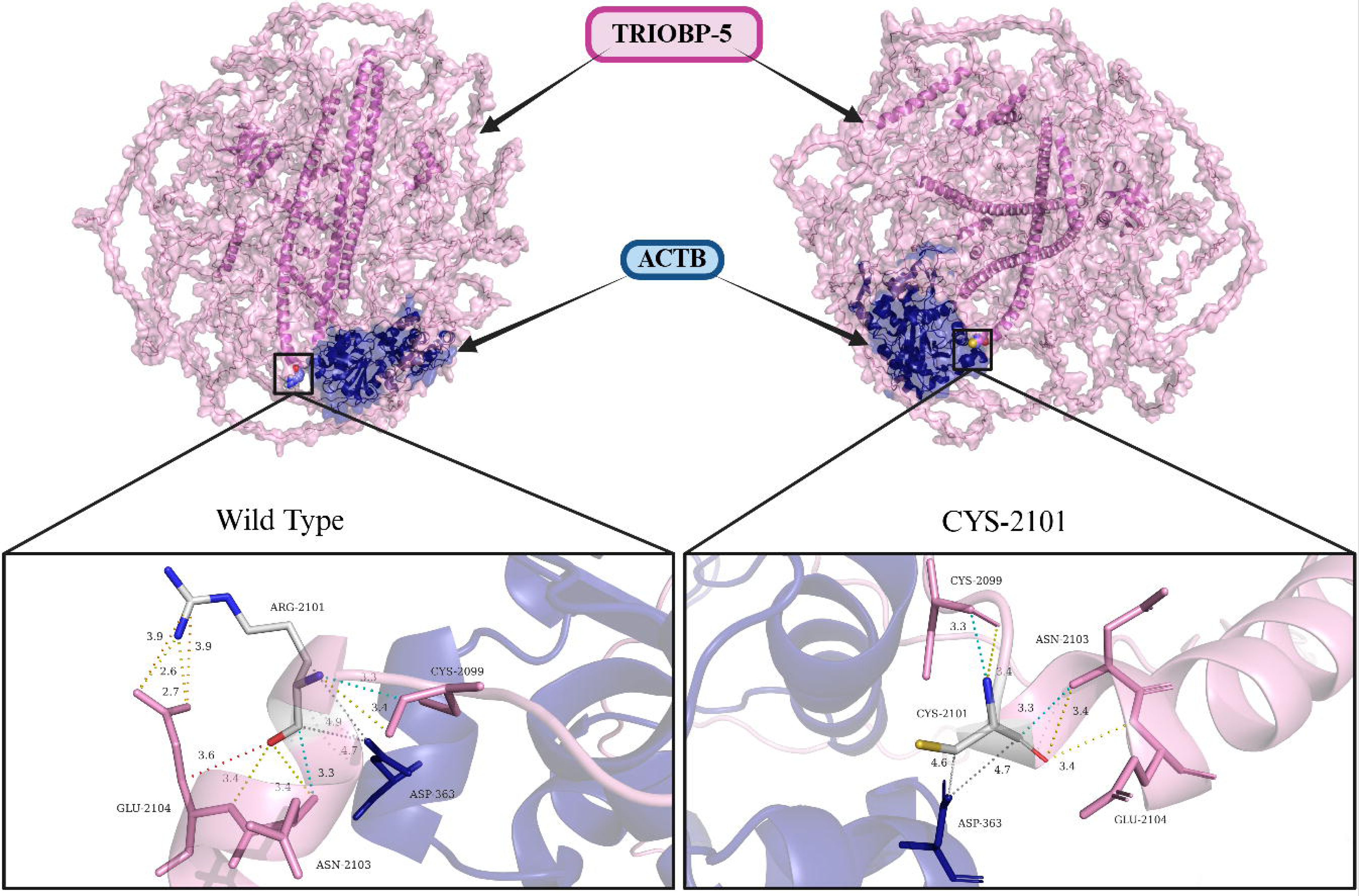
Wild type (left) and mutant (right) *TRIOBP*-5 and ACTB interaction model, highlighting atomic interaction between wild type (left) and mutant (right) *TRIOBP*-5 p.2101 and ACTB p.D363

Similarly, *TRIOBP*-6 residues p.R2273 and p.C2273 exhibited no interactions with either G-actin or F-actin.

Interestingly, the dimer structure of *TRIOBP*-5 docked with β-actin revealed significant change on the predicted behaviour of their interaction. Both wt and mutated amino acids lose and gain contacts with neighbouring residues when compared to the monomer docking. Amino acid interactions between *TRIOBP*-5 (p.2101) and β-actin (p.363) occur only in one of *TRIOBP*-5 chains (chain C), rather than in both (chains A and C) (**Figures A21-A22**). DynaMut2 predicted that the change of arginine for cytosine stabilise chain A of the molecule in 0.78 kcal^-1^ and chain C, in 0.67 kcal^-1^.

## Identification of *TRIOBP* Protein in the Cochlear and Vestibular Hair Cells

*TRIOBP* is expressed in the stereocilia rootlets of the cochlear hair cells (**Fig 5**). As expected stereocilia rootlets of the cochlear hair cells also stained for *TRIOBP.* Similarly, stereocilia rootlets of the utricle from the vestibular system also stained for *TRIOBP* protein.

**Figure 5.**
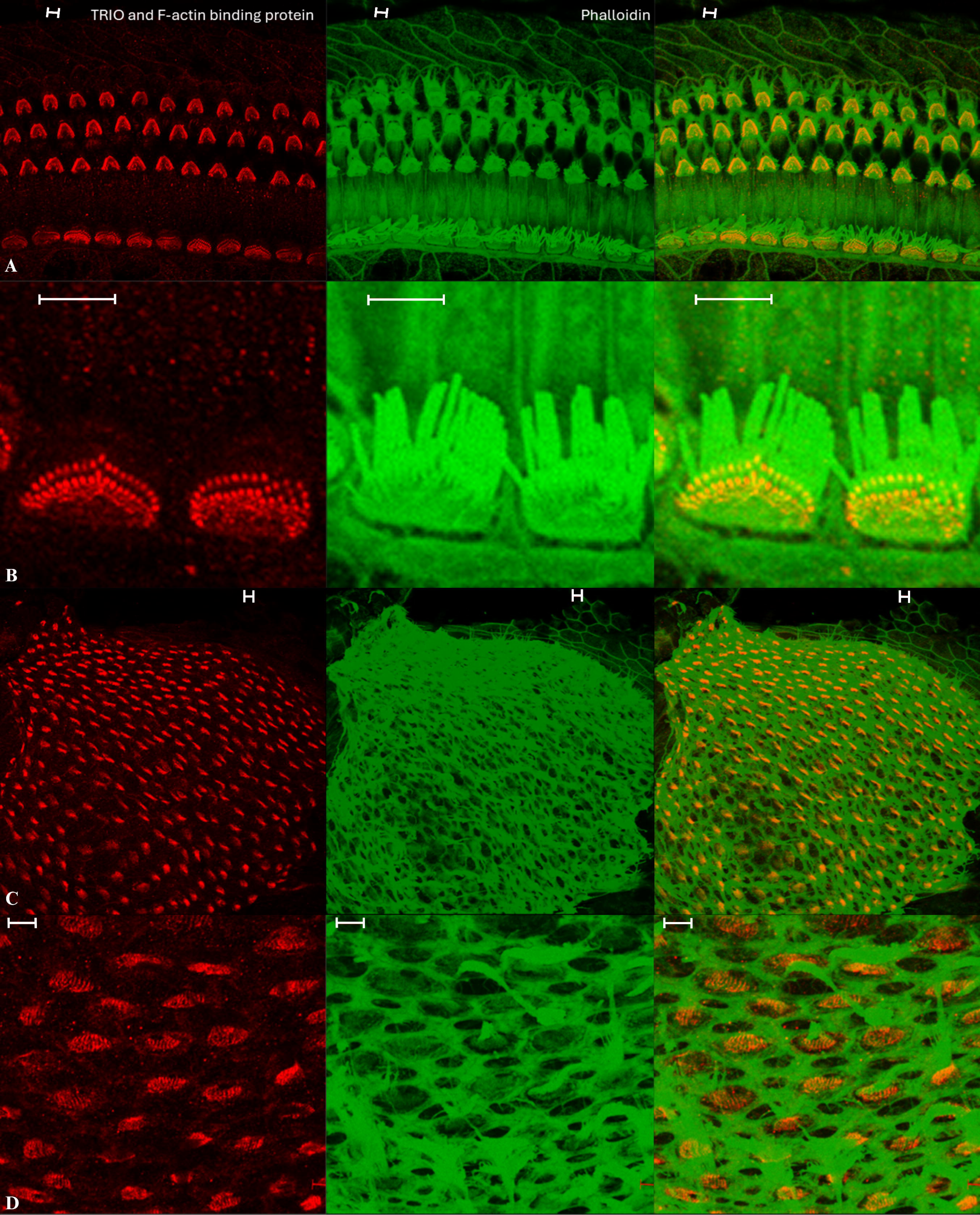
A) *TRIOBP* (TRIO and F actin binding protein) is expressed in the stereocilia rootlets of wild type (wt) mouse inner hair cells (IHC) and outer hair cells (OHC). B. *TRIOBP* in outer hair cell stereocilia rotlets. Immunofluorescence analysis of *TRIOBP* in P14 whole- mount mouse inner ears stained with *TRIOBP* and phalloidin to visualize filamentous actin. Scale bars in panels, 5μm. C and D) *TRIOBP* is expressed in the stereocilia rootlets of wt mouse vestibular hair cells of the utricle. Immunofluorescence analysis of *TRIOBP* in P14 whole- mount mouse inner ears stained with *TRIOBP* and phalloidin to visualize filamentous actin. Scale bars in panels, 5μm.

*TRIOBP* expression in the utricle suggested its expression to be more prominent in the vestibular system. To end this, we stained Cristae Ampullaris of the semicircular canal. Interestingly, we found expression of *TRIOBP* in the rootlet of the hair cells (**Fig 6**).

**Figure 6.** *TRIOBP* (TRIO and F actin binding protein) is expressed in the stereocilia rootlets of wild type (wt) of cristae ampullaris. Immunofluorescence analysis of *TRIOBP* in P14 whole- mount mouse inner ears stained with *TRIOBP* and phalloidin to visualize filamentous actin. Inlet shows higher magnification of the stereocilia rootlets expressing *TRIOBP*. Scale bars in panels, 5μm.

## Discussion

In our study, we analysed exome sequencing data from individuals with MD who had a history of occupational noise exposure, with the aim of identifying rare variants that may contribute to the MD noise-induce MD phenotype. Notably, we identified a heterozygous missense variant in *TRIOPB* (chr22:37769343C>T) in two of the 34 individuals of European ancestry with a history of occupational noise exposure. This variant is a strong candidate for contributing to the auditory phenotype observed in this cohort. Pathogenic variants in *TRIOBP* have been associated with DFNB28 across multiple populations.^19^ The identification of this rare variant in MD individuals with occupational noise exposure supports the hypothesis that rare mutations in *TRIOBP*, in combination with environmental factors such as noise exposure, may contribute to change the stiffness of hair cell stereocilia and development of MD.

Experimental studies have shown that the *TRIOBP*-5 isoform, specifically binds and crosslinks F-actin through its N-terminal repeats. This has been demonstrated through co-sedimentation assays and immunofluorescence in inner ear stereocilia rootlets, near the apical surface.^18,57^ Actin exists in monomeric (G-actin) and filamentous (F-actin) states, with β-actin representing one of six conserved mammalian isoforms predominantly expressed in non-muscle cells.^58,59^ The polymerisation of ATP-bound G-actin into double-helical F-actin filaments provides mechanical support for cellular structures.^60^ This interaction between *TRIOBP*-Actin is functionally significant, as pathogenic *TRIOBP*-5 mutations (e.g., p.R785W) disrupt F-actin bundling and cause DFNB28 deafness.^61^ Additionally, IMPC data indicate that *TRIOBP*-knockout exhibit severe hearing loss across all frequencies.^52^ However, the IMPC does not report whether these mice display any vestibular phenotype.

Furthermore, we found a mutation in the *LGALS8-AS1*(chr1:236524482A>G) lncRNA. *LGALS8-AS1* has been found to regulate the expression of fascin actin-binding protein 1 (*FSCN1* gene) by targeting microRNA (miRNA) miR-885-3p.^62^ While fascin actin-binding protein 1 has not been described in the stereocilia rootlet, it has been found to bind to the F-actin in the stereocilia core.^63,64^ TRIO and F-actin-binding protein (*TRIOBP* gene) isoform 5 function in the inner ear is to reinforce F-actin protein bundles in the stereocilia rootlet to maintain rigidity to allow for mechanotransduction.^57^

We retrieved ATAC-seq DA peak data and observed significant downregulation of two UTR regions of the transcription factor *RFX3* in MD cases compared to controls. A previous study has shown that *RFX3* regulates *TRIOBP* expression in mice inner ear hair cells by binding to an intronic cis-regulatory element, acting potentially as either an activator or a repressor.^56^ Consistent with this, our data show that the *TRIOBP* DA peaks are also downregulated in the same patient cohort, suggesting that the downregulation of *RFX3* 3’ UTR regions could lead to reduced *TRIOBP* transcription in MD case, potentially contributing to weaker stereocilia rootlets and contributing to hearing loss, particularly when exposed to occupational noise.

While *TRIOBP*-6 is the longest isoform, studies are usually performed in *TRIOBP*-5 as it is expressed in both humans and murine species. The function of *TRIOBP*-6 is hypothesised to be similar to the one of *TRIOBP*-5, as they include the same domains.^21^ Our results also predict an interaction between *TRIOBP*-6 and *NIN*. The ninein protein (*NIN* gene) shows dynamic localisation during centriole migration in hair cells. In early stages (Phase I), it is concentrated around the centrally positioned mother and daughter centrioles (MC, DC respectively). As the DC moves towards the cell periphery (Phase II), ninein distribution broadens, suggesting a role in centriole positioning and hair cell polarity.^65^ Furthermore, the IMPC has established a relationship between high frequency hearing loss and *NIN*-knockout.^51^

Studies in mice have demonstrated that the *TRIOBP*-5 isoform interacts with ankyrin repeat domain-containing protein 24, encoded by the *ANKRD24* gene. *ANKRD24* functions to organize the stereocilia rootlet by encapsulating *TRIOBP*-5, which subsequently bundles F-actin within the rootlet structure, thereby providing additional stability.^66^ Furthermore, a recent study identified a burden of rare variants in *ANKRD24* and *NIN* genes among both sporadic and familial MD patients, suggesting that mutations in stereocilia rootlet-associated genes may contribute to the pathogenesis of MD phenotypes.^8^

By using confocal microscopy and immunofluorescent antibodies for *TRIOBP* protein and ninein, we have shown that these proteins are expressed in the mouse auditory and vestibular hair cells stereocilia, with *TRIOBP* located at the base of each stereocilia, in both inner and outer hair cells, but also in vestibular hair cells.

Most genes previously linked in MD, such as *OTOG*, *MYO7A* and *TECTA*, encode proteins localised to structures like the stereocilia’s tip links, horizontal top connectors, and the tectorial membrane.^4,6,67^ Additionally, a rare haplotype of *GJD3* has been linked to sporadic and familial MD, with its protein product enriched in the base of inner and outer hair cells.^5^ These findings suggest that disruption of the structural integrity of stereocilia may contribute to the audiovestibular phenotype in MD. Our findings expand this hypothesis to *TRIOBP*, a gene essential for stereocilia rootlet structure, as a potential contributor to the MD pa in noise-induce hearing loss. Further studies are needed to validate the functional impact of this variant and to explore gene-environment interactions, including noise exposure in the pathophysiology of MD.

Our study has some limitations. First, the sample size is limited since the identification of patients with MD and occupational noise exposure is not common. Second, most of the genetic studies in MD are limited to European descendent population and data with different ancestry are needed to validate these variants in *TRIOBP*. Finally, *TRIOPB* protein has a confirmed interaction with β-actin and our candidate mutation interfere this interaction, we could not demonstrate a physical interaction between *TRIOBP* protein and ninein.

## Conclusions

An ultra-rare variant in the *TRIOBP* gene was found in several unrelated MD individuals with history of occupational noise exposure. Computational modelling revealed that the variant modified atomic bonds between *TRIOBP* and *ACTB* proteins, leading to increased stability of the dimer. These findings support that rare mutations in *TRIOBP*, in combination with environmental factors such as noise exposure, may contribute to change the stiffness of hair cell stereocilia and development of MD.

## Data Availability

Data Availability
TRIOBP chr22:37769343C>T variant has been deposited in ClinVar under accession number XXXXX.

https://www.ncbi.nlm.nih.gov/clinvar/

## Author Contributions

P.C.G -

G.B.B –

Y.Z –

P.T.N.P –

K.B.L -

E.L.S –

I.S.S -

A.S.V -

J.A.L.E – Conceptualization, design, data analysis, writing first draft, final draft.

## Acknowledgement

We acknowledge the patients for donating blood for this study.

## Data Availability

*TRIOBP* chr22:37769343C>T variant has been deposited in ClinVar under accession number XXXXX.

## Abbreviations

Menière’s Disease (MD), Sensorineural hearing loss (SNHL), Noise-induced hearing loss (NIHL), Stria vascularis (SV), Marginal cells (MC), Intimidate cells (IMC), Basal cells (BC), Non-Finnish European (NFE), Long non-coding RNA (lncRNA), Exonic Splicing Enhancer (ESE), Exonic Splicing Silencers (ESS), Intronic Splicing Enhancers (ISE), Intronic Splicing Silencers (ISS), American College of Medical Genetics and Genomics(ACMG)

Appendix A – Supplementary Data

