## Supplementary Data for "A Rare Variant in *TRIOBP* Linked to Occupational Noise Exposure in Meniere Disease"

**List of Tables and Figures**

**Table A1.-** *TRIOBP* differentially methylated CpG sites in MD patients.

| Chromosome | Position | Gene Name | Average Methylation | Min Value | Max Value |
|---|---|---|---|---|---|
| chr22 | 37711565 | *TRIOBP* | 97.14 | 84.85 | 100 |
| chr22 | 37711573 | *TRIOBP* | 94.13 | 76.47 | 100 |
| chr22 | 37711631 | *TRIOBP* | 69.63 | 31.25 | 100 |
| chr22 | 37711741 | *TRIOBP* | 97.75 | 87.50 | 100 |
| chr22 | 37711748 | *TRIOBP* | 91.58 | 77.78 | 100 |
| chr22 | 37729697 | *TRIOBP* | 95.41 | 84.62 | 100 |

**Table A2.-** Differentially accessible peaks in *TRIOBP* genes on single-cell ATACseq MD patients.

| Region | avg_log2FC | *p*_val_adj | gene_name | gene_biotype | type | distance |
|---|---|---|---|---|---|---|
| **MD - External Controls** | | | | | | |
| chr22-37696386-37697412 | -1.50 | $2.97x10^{-04}$ | *TRIOBP* | protein_coding | utr | 0 |
| chr22-37739182-37741840 | 0.13 | 1 | *TRIOBP* | protein_coding | cds | 0 |
| chr22-37745329-37747609 | -1.01 | 1 | *TRIOBP* | protein_coding | cds | 0 |
| **MD - Internal Controls** | | | | | | |
| chr22-37745682-37746612 | -0.48 | 1 | *TRIOBP* | protein_coding | cds | 0 |

**Table A3.-** Splice site prediction for *TRIOBP* chr22:37769343C>T variant in transcripts ENST00000403663 and ENST00000644935.

| | |
|---|---|
| **Algorithme/Matrix** | HSF Donor site (matrix GT) |
| **Position** | chr22:37769339 |
| **Sequences** | CGGGCGCAG>CGGGTGCAG |
| **Variation** | 44.03>71.17 (61.64%) |
| **Signal** | New Donor splice site |
| **Interpretation** | Activation of a cryptic Donor site. Potential alteration of splicing |
| **Associated transcripts** | ENST00000403663; ENST00000644935 |

**Table A4.-** Creation and deletion of Exonic Splicing Enhancers (ESE) and Exonic Splicing Silencers (ESS) regulatory elements in *TRIOBP* chr22:37769343C>T variant.

| Regulatory Elements | Name | Position | Sequence | Status |
|---|---|---|---|---|
| ESE | ESE_9G8 | chr22:37769338 | GCGGGC | Site Broken |
| ESE | ESE_ASFB | chr22:37769339 | CGGGCGC | Site Broken |
| ESS | IIE | chr22:37769340 | GGGTGC | Site Created |
| ESS | Sironi_motif2 | chr22:37769341 | GGTGCAG | Site Created |
| ESE | PESE | chr22:37769341 | GGCGCAGC | Site Broken |
| ESE | ESE_ASF | chr22:37769343 | CGCAGCA | Site Broken |
| ESE | ESE_ASFB | chr22:37769343 | CGCAGCA | Site Broken |

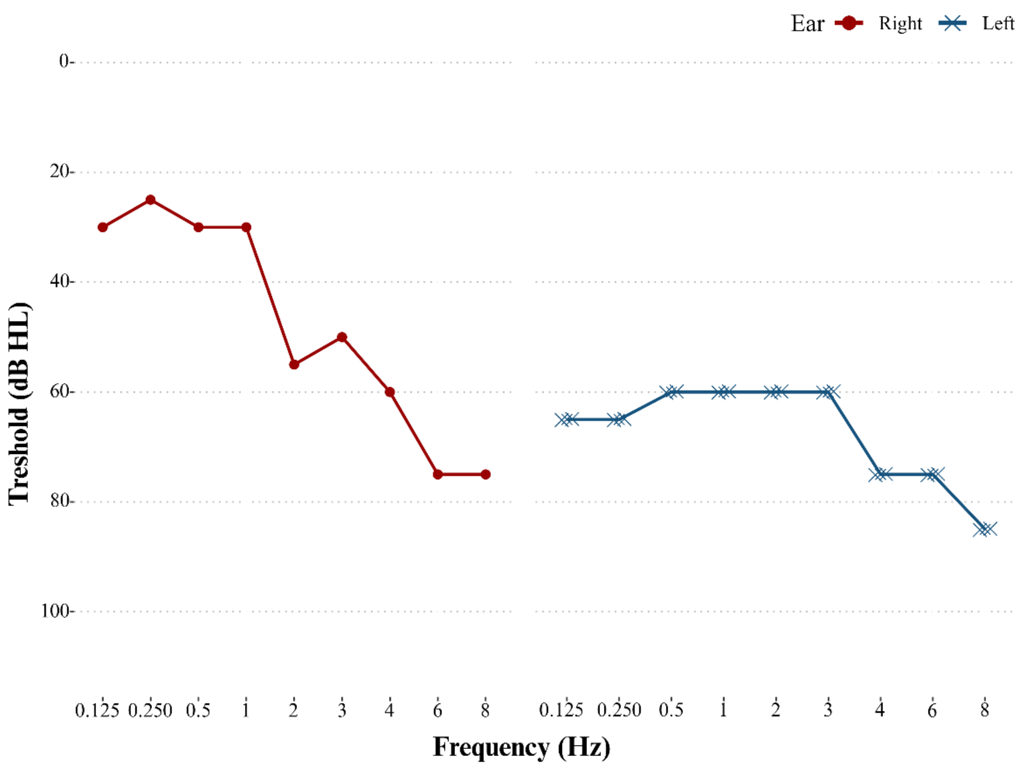

**Figure A1.-** Meniere Disease individual 3 with occupational noise exposure. Audiogram showing air-conducted pure tone hearing thresholds in their seventh decade of life with bilateral high-frequency hearing loss and SNHL involving all frequencies in the left ear. Right ear is represented in red, left ear in blue.

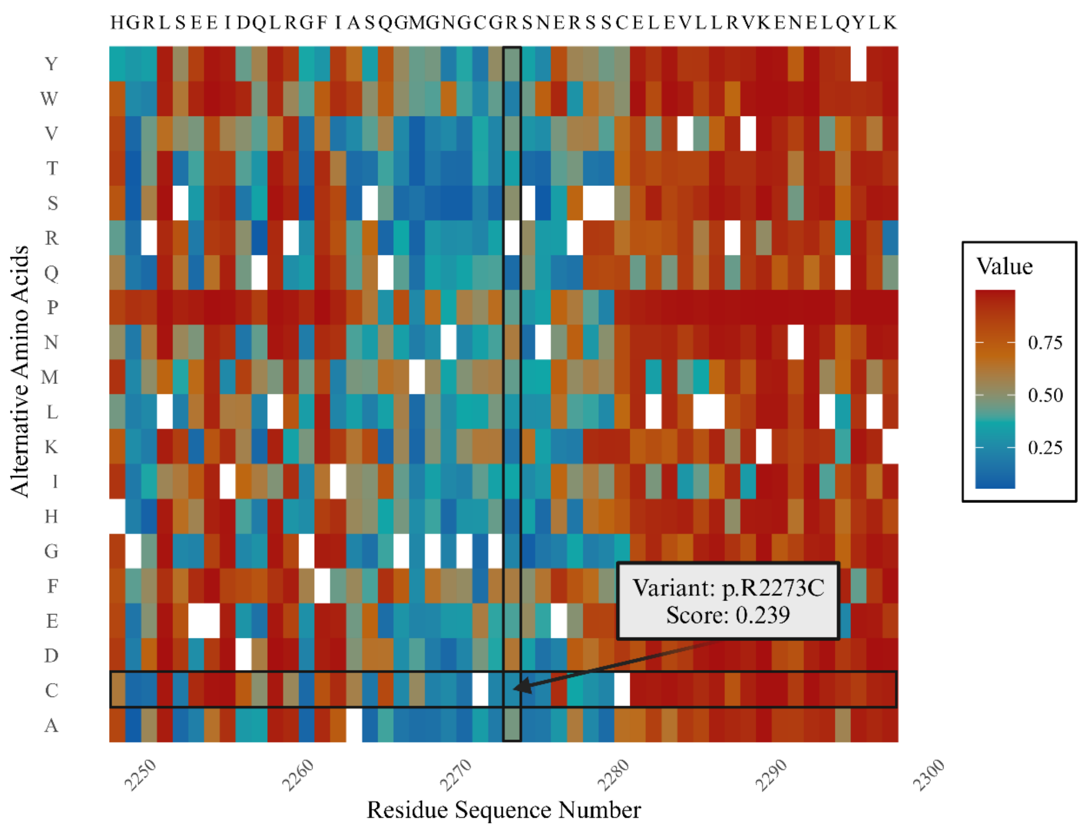

**Figure A2.-** *TRIOBP* p.R2273C (chr22:37769343C>T) pathogenicity heatmap. AlphaMissense scores *TRIOBP* variant p.R2273C as likely being with a value of 0.239.

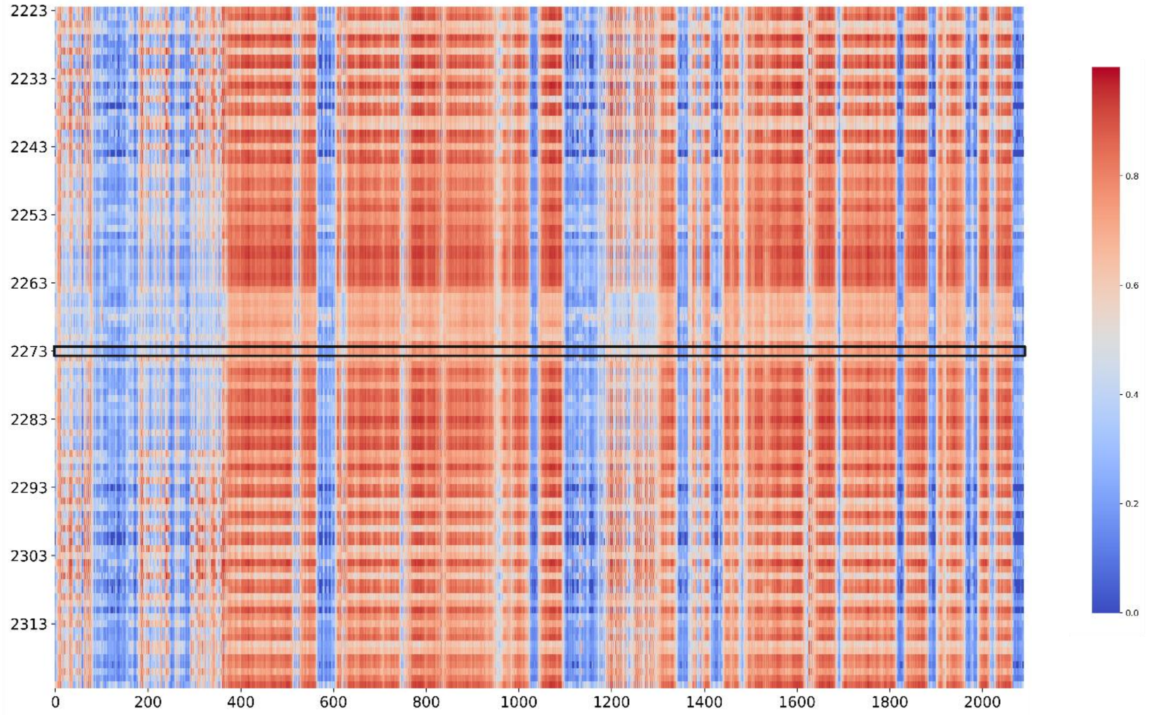

**Figure A3.-** Docknet prediction of *TRIOBP*-6 p.R2273 and *NIN*

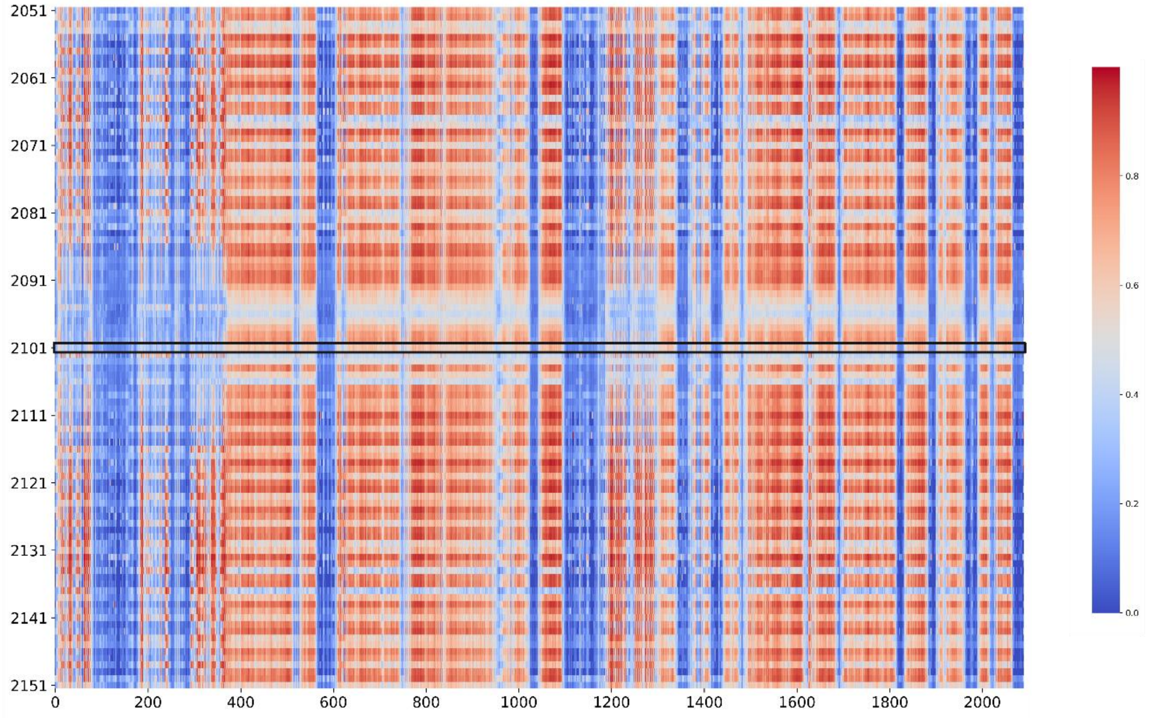

**Figure A4.-** Docknet prediction of *TRIOBP*-5 p.R2101 and *NIN*

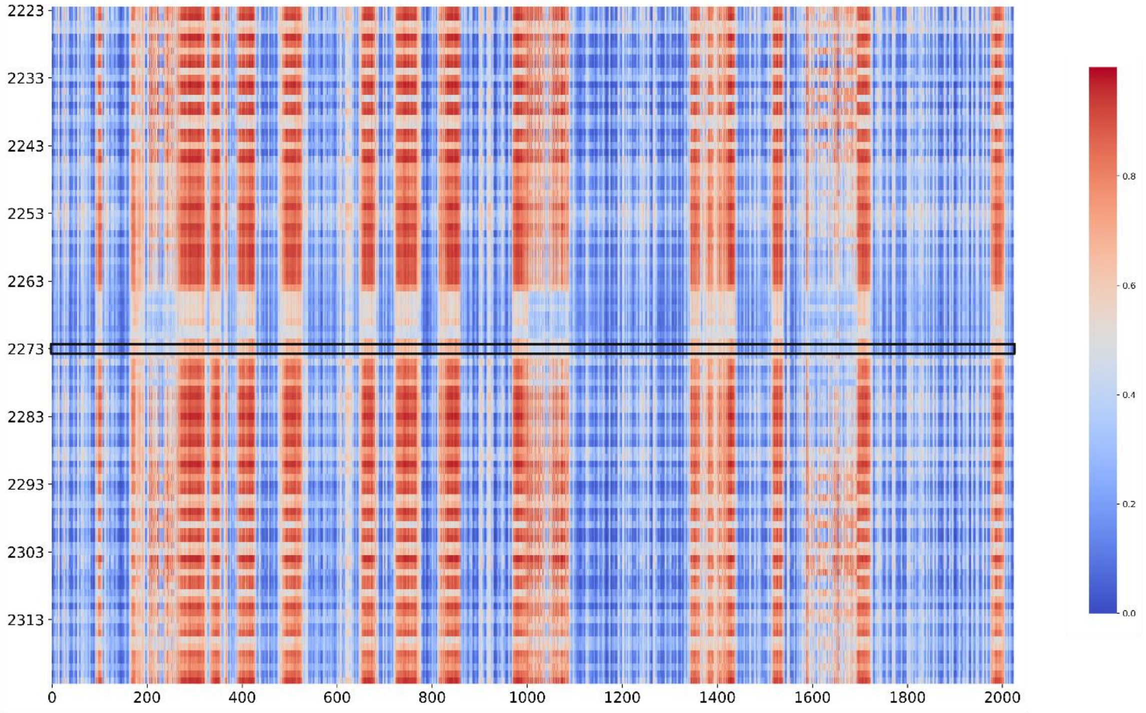

**Figure A5.-** Docknet prediction of *TRIOBP*-6 p.R2273 and *PCM1*

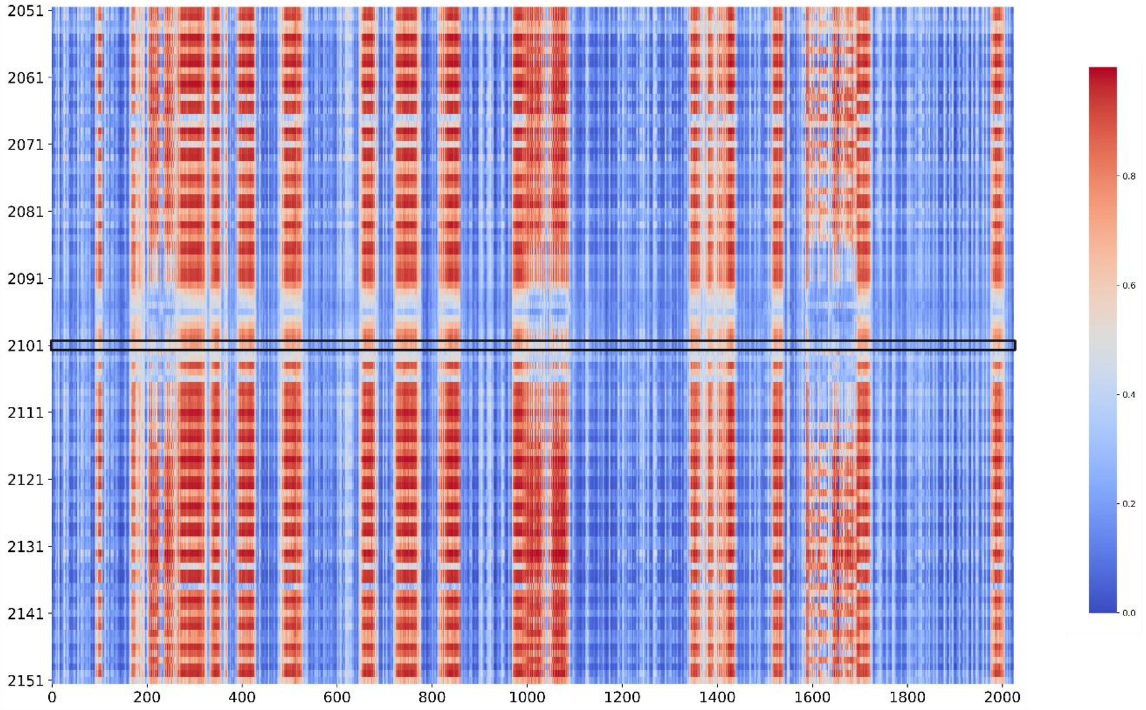

**Figure A6.-** Docknet prediction of *TRIOBP*-5 p.R2101 and *PCM1*

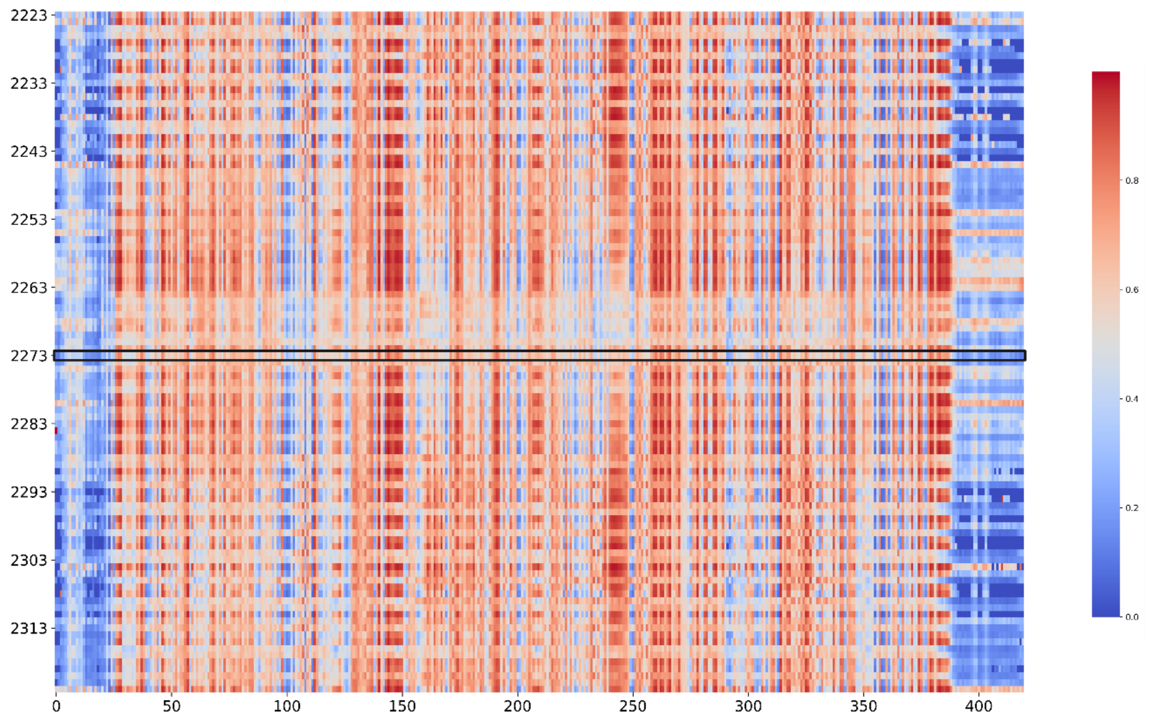

**Figure A7.-** Docknet prediction of *TRIOBP*-6 p.R2273 and *GSK3B*

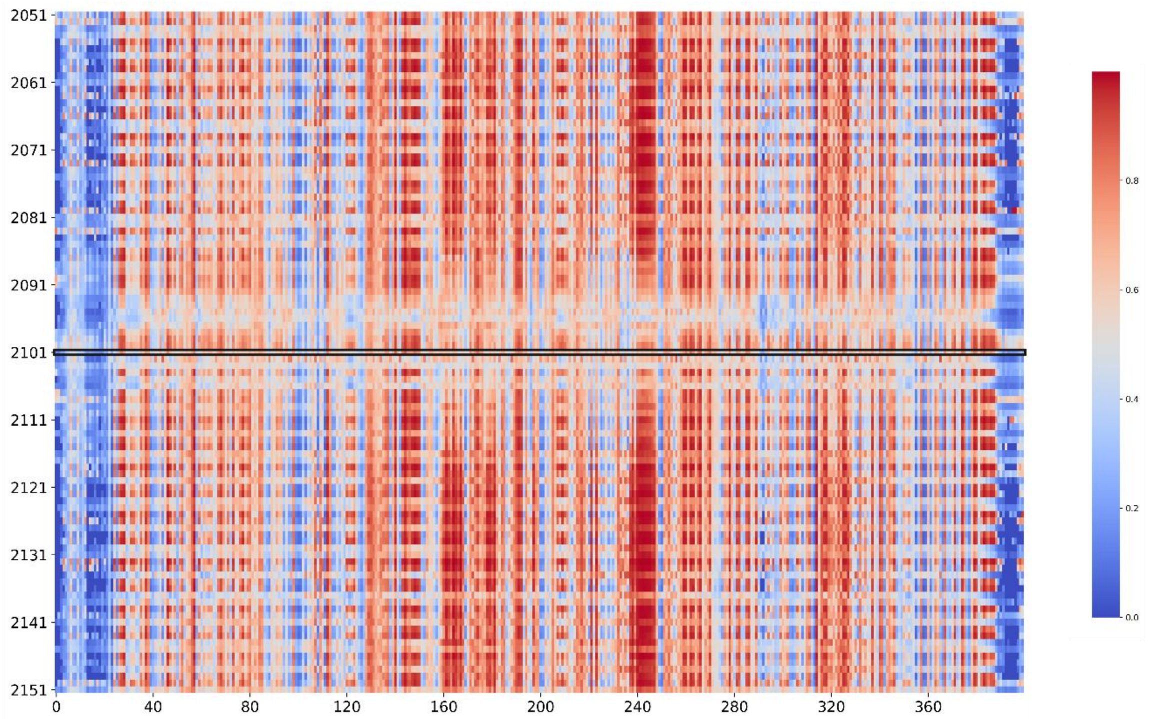

**Figure A8.-** Docknet prediction of *TRIOBP*-5 p.R2101 and *GSK3B*

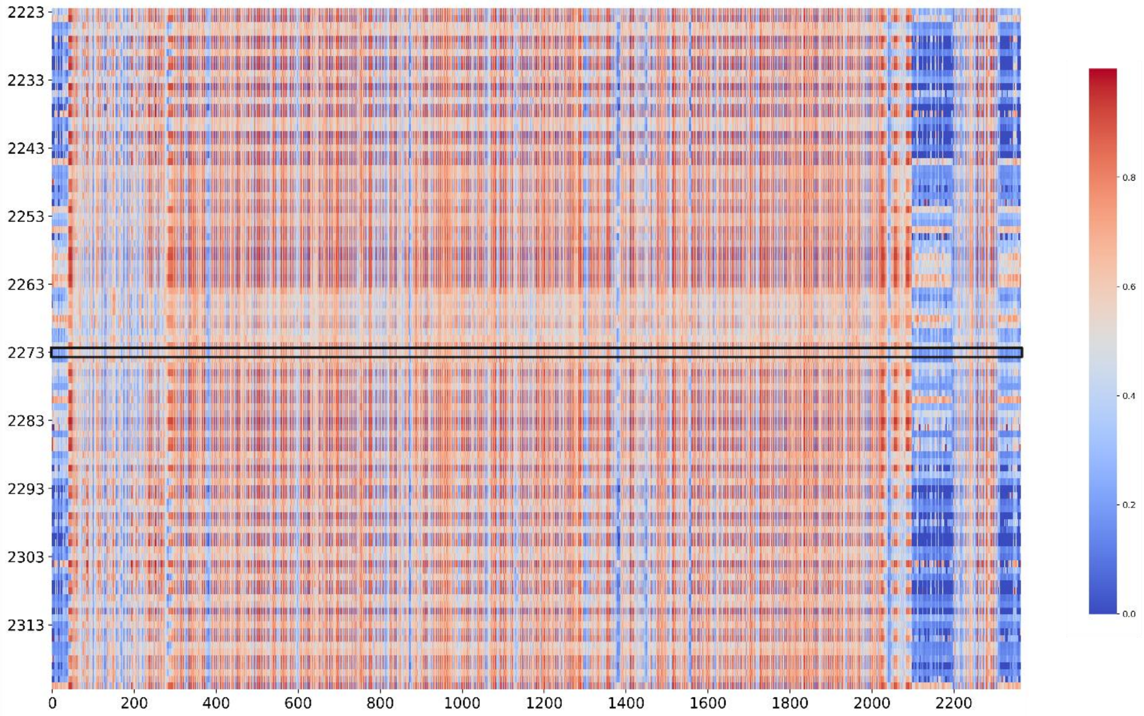

**Figure A9.-** Docknet prediction of *TRIOBP*-6 p.R2273 and *SPTBN1*

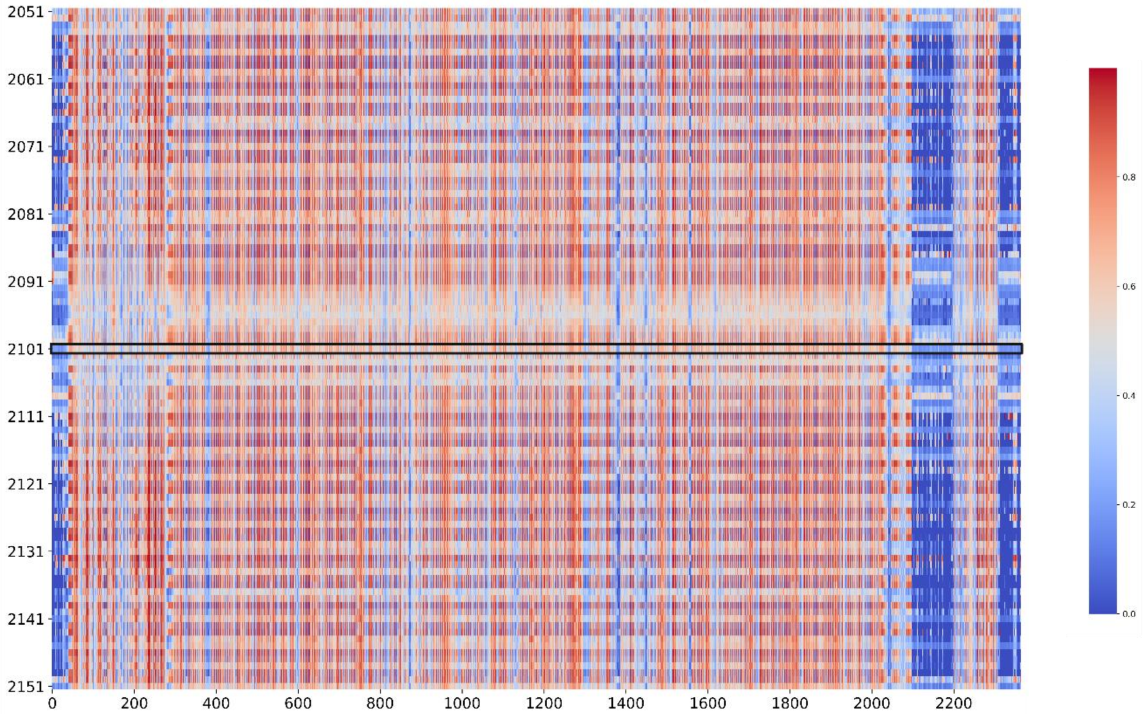

**Figure A10.-** Docknet prediction of *TRIOBP*-5 p.R2101 and *SPTBN1*

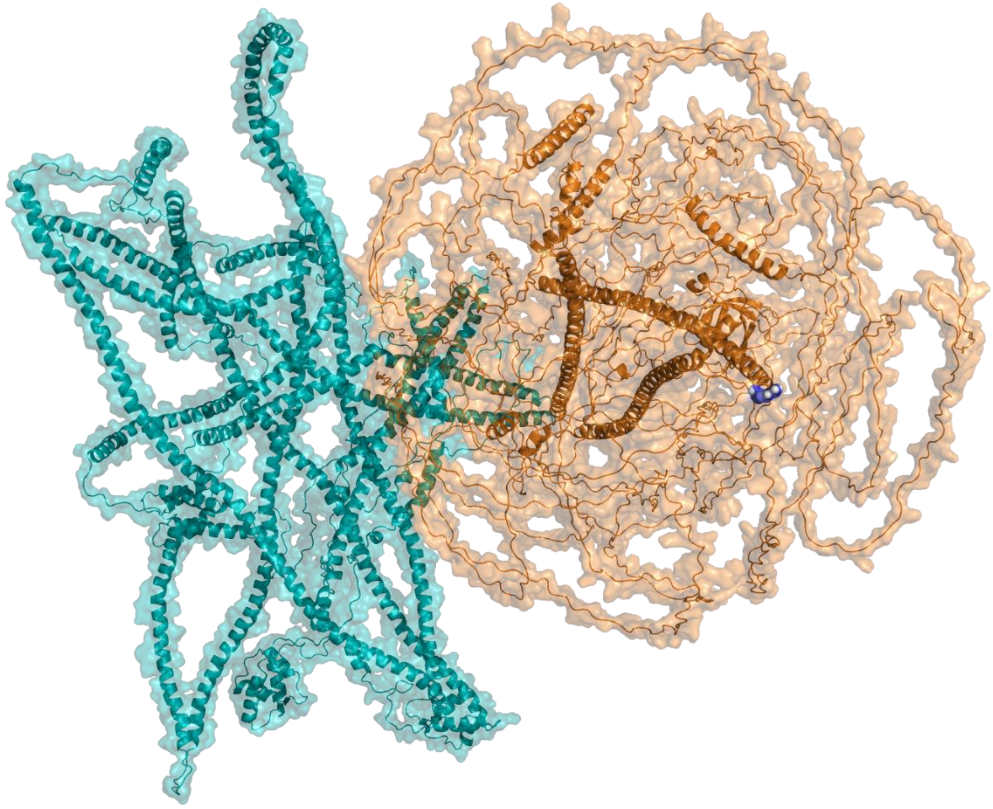

**Figure A11.-** Docked model between wt *TRIOBP*-6 (orange) and *NIN* (cyan). Wt p.2273R was not predicted to interact with *NIN*.

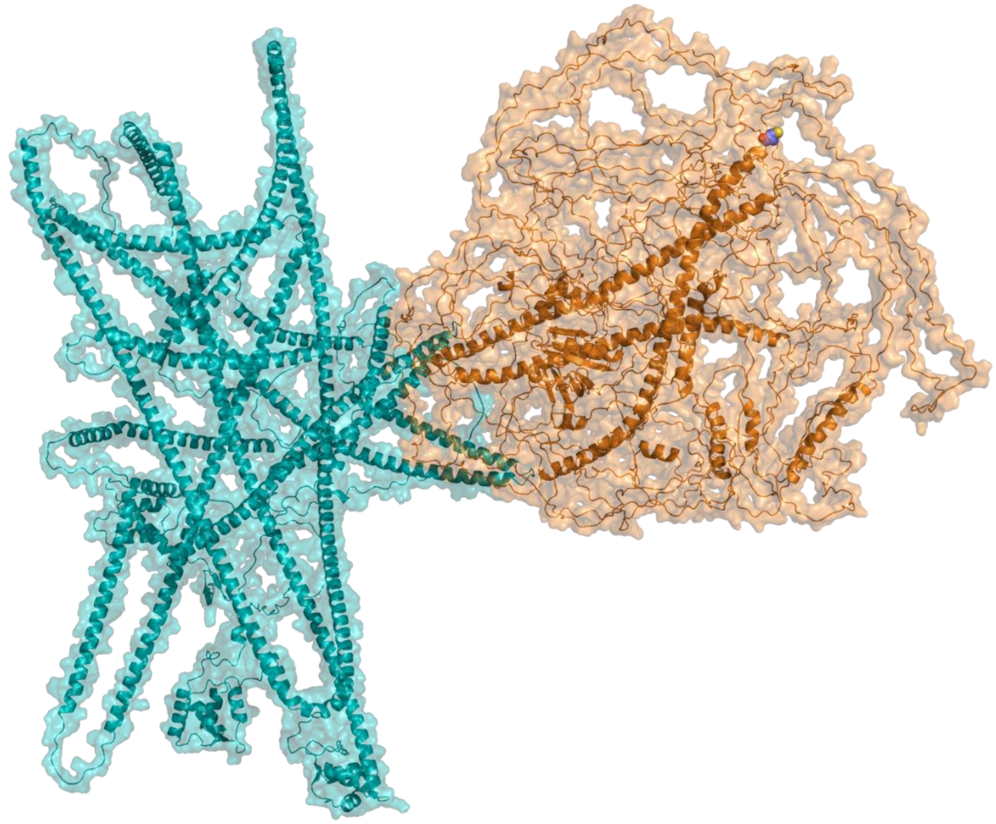

**Figure A12.-** Docked model between mt *TRIOBP*-6 (orange) and *NIN* (cyan). Variant p.2273C was not predicted to interact with *NIN*.

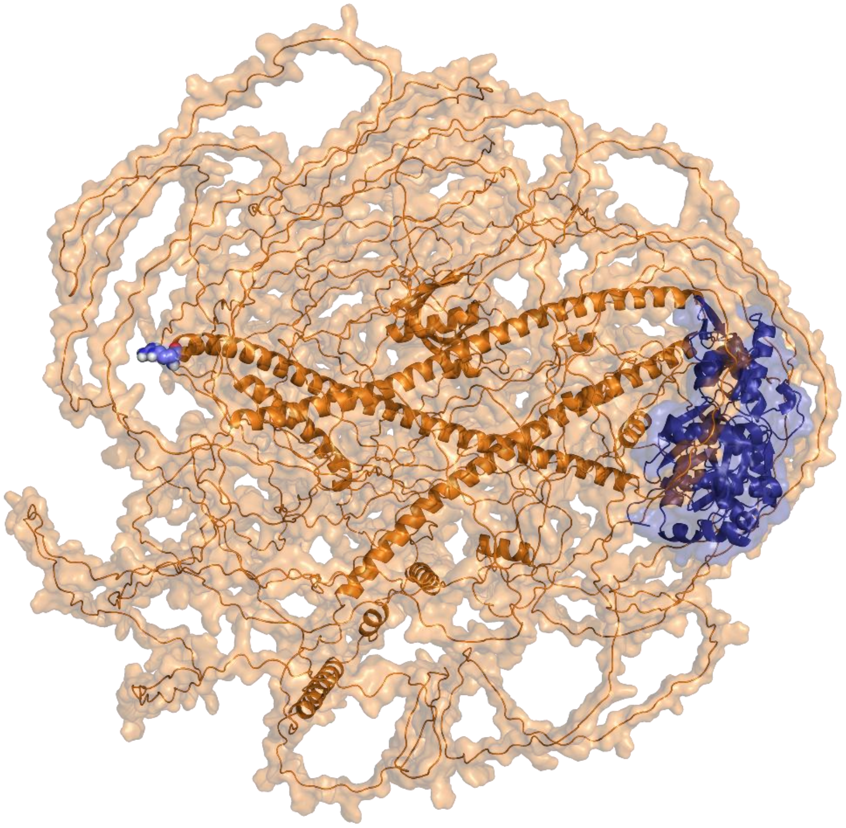

**Figure A13.-** Docked model between wt *TRIOBP*-6 (orange) and *ACTB* (blue). Variant p.2273R was not predicted to interact with *ACTB*.

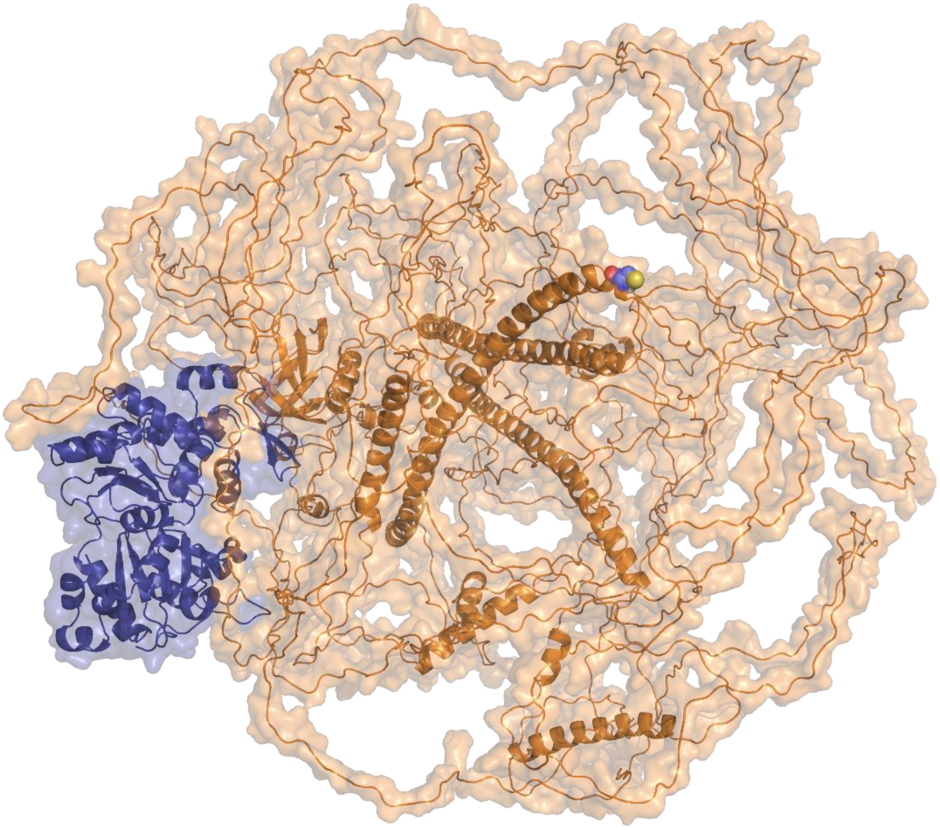

**Figure A14.-** Docked model between mt *TRIOBP*-6 (orange) and *ACTB* (blue). Variant p.2273C was not predicted to interact with *ACTB*.

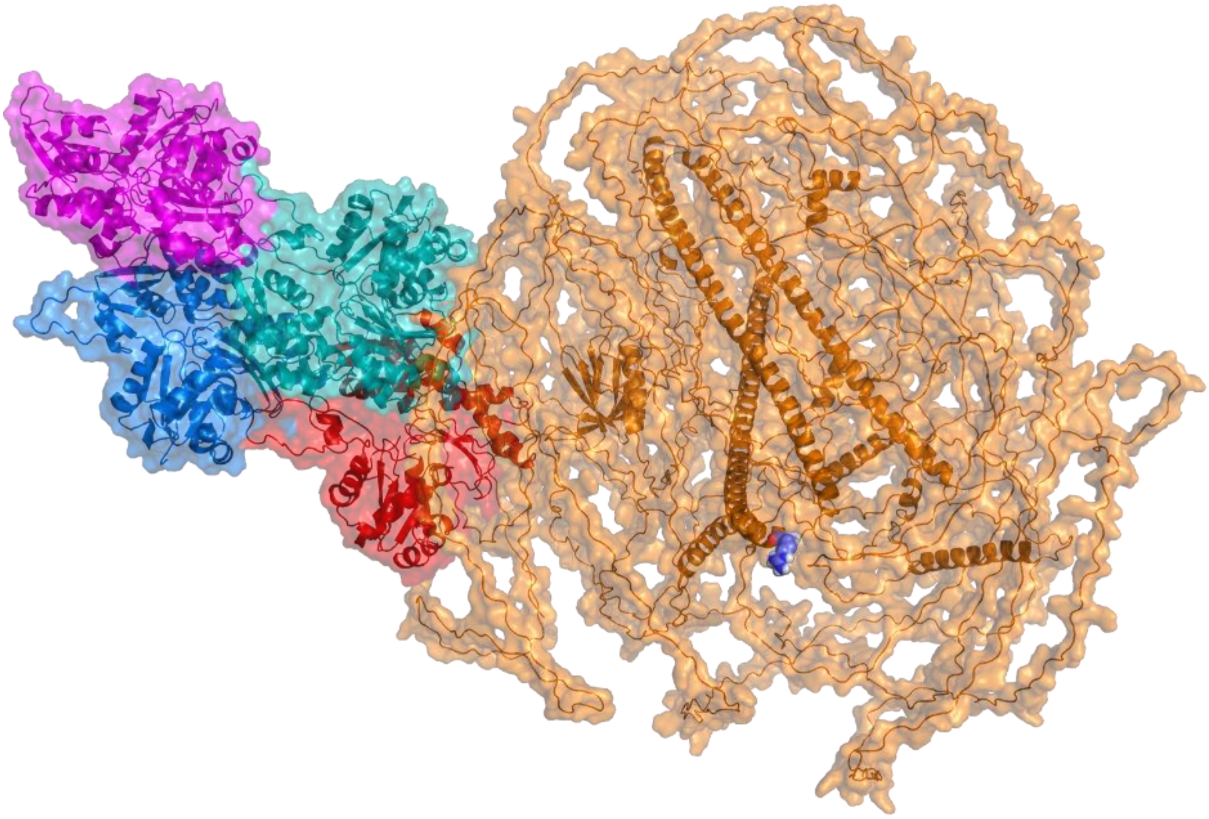

**Figure A15.-** Docked model between wt *TRIOBP*-6 (orange) and F-Actin (magenta, cyan, blue and red). Variant p.2273R was not predicted to interact with F-Actin.

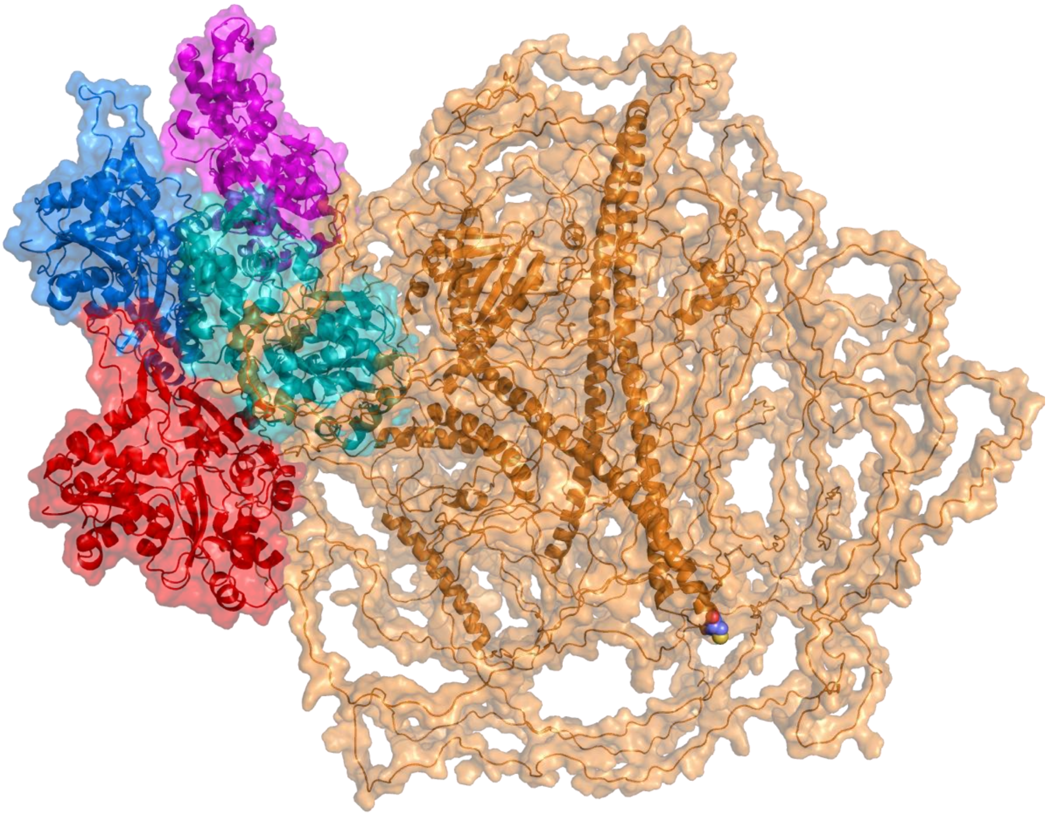

**Figure A16.-** Docked model between mt *TRIOBP*-6 (orange) and F-Actin (magenta, cyan, blue and red). Variant p.2273C was not predicted to interact with F-Actin.

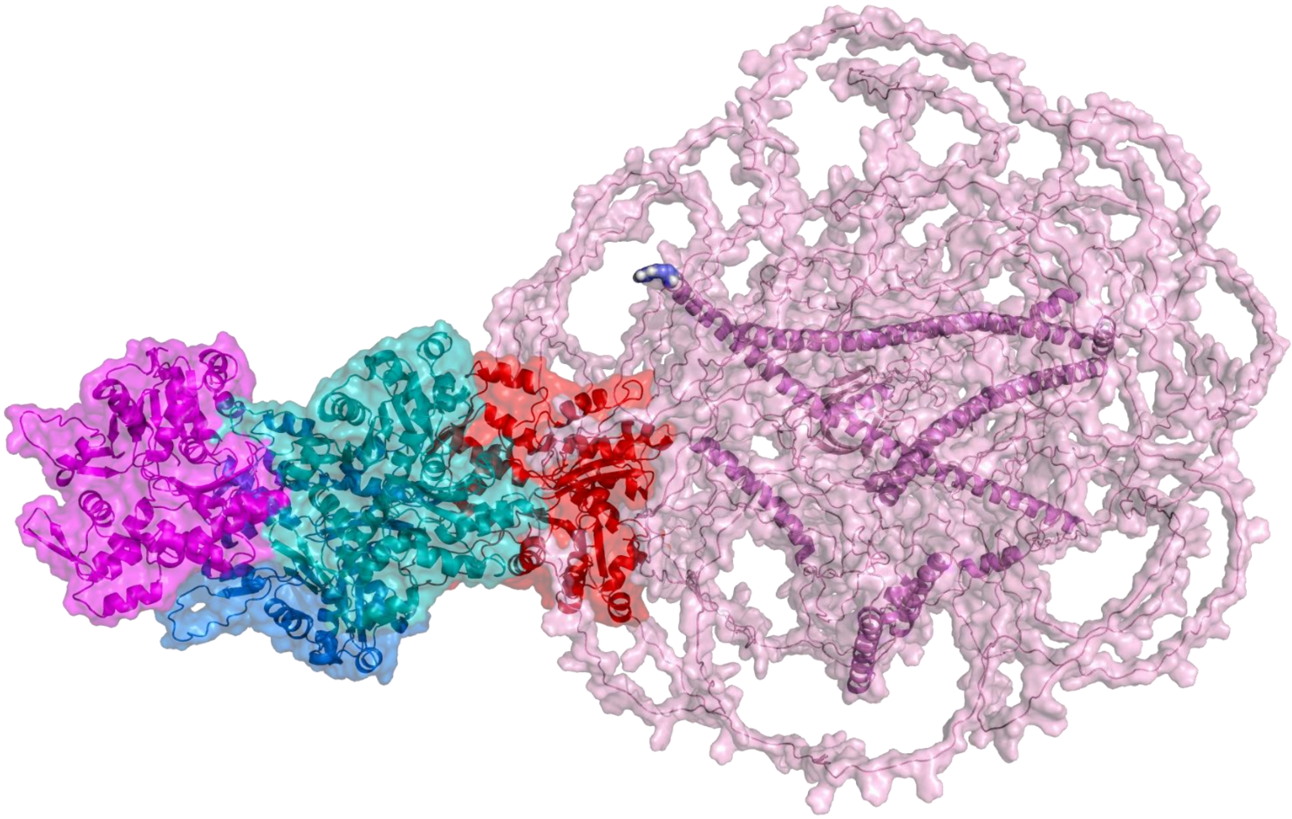

**Figure A17.-** Docked model between wt *TRIOBP*-5 (orange) and F-Actin (magenta, cyan, blue and red). Variant p.2273R was not predicted to interact with F-Actin.

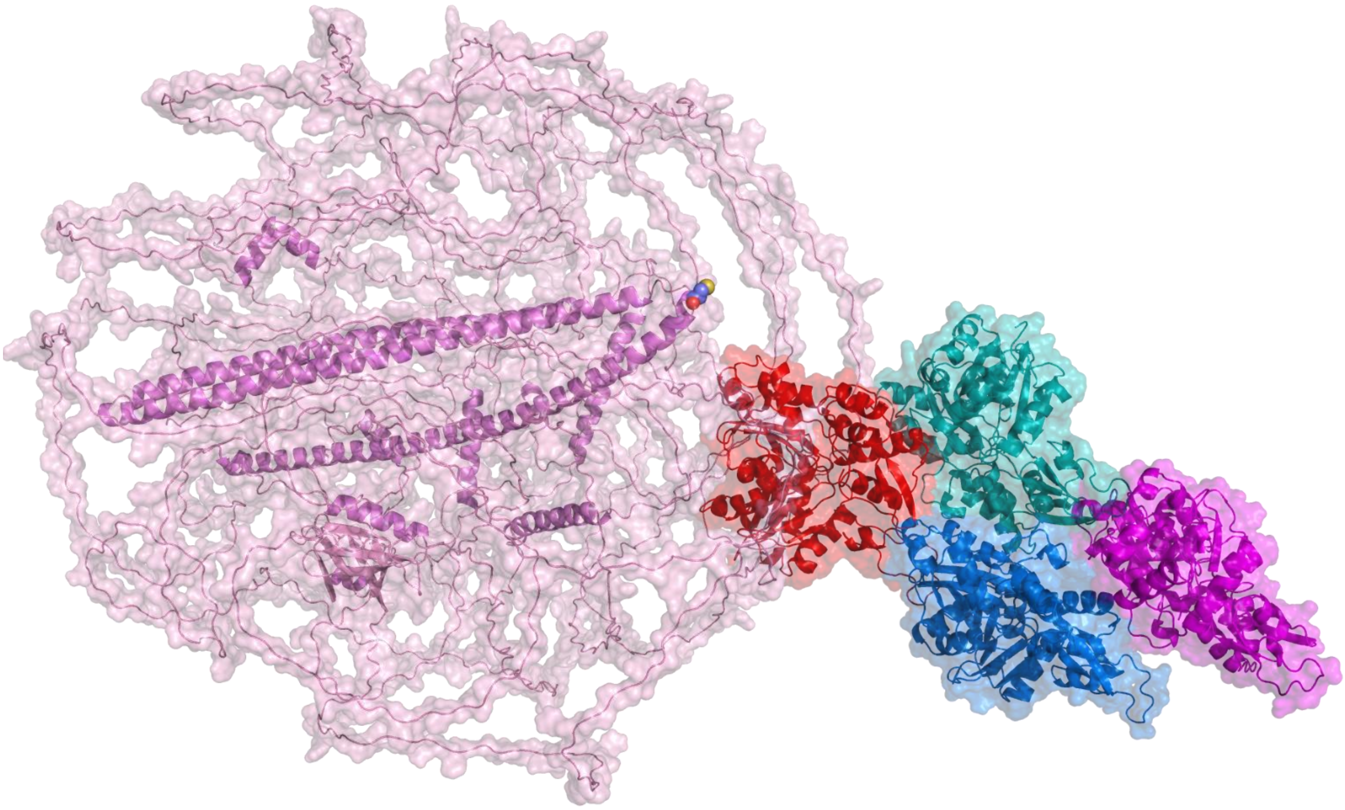

**Figure A18.-** Docked model between mt *TRIOBP*-5 (orange) and F-Actin (magenta, cyan, blue and red). Variant p.2273C was not predicted to interact with F-Actin.

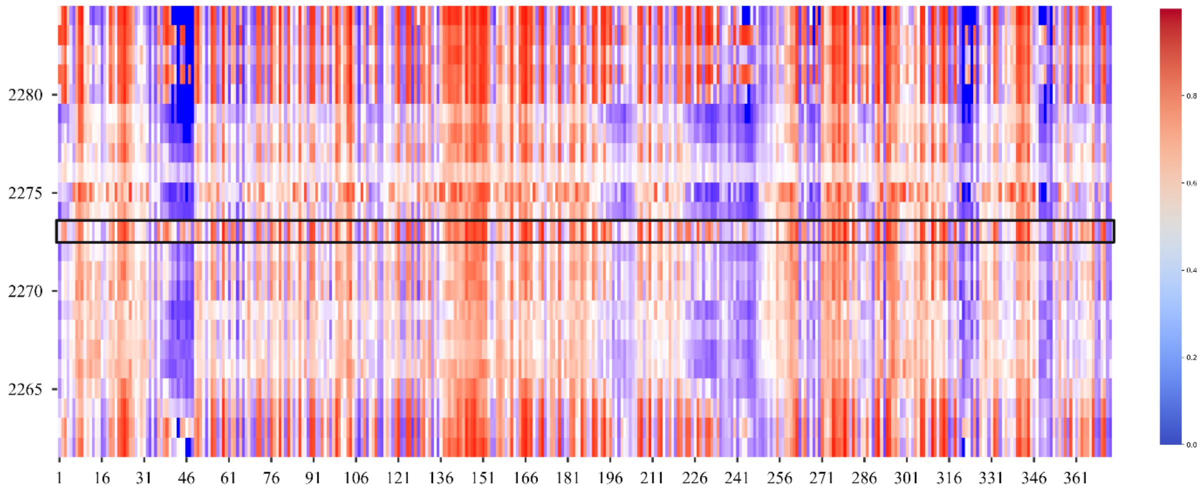

**Figure A19.-** Docknet prediction of *TRIOBP*-6 p.R2273 and *ACTB*

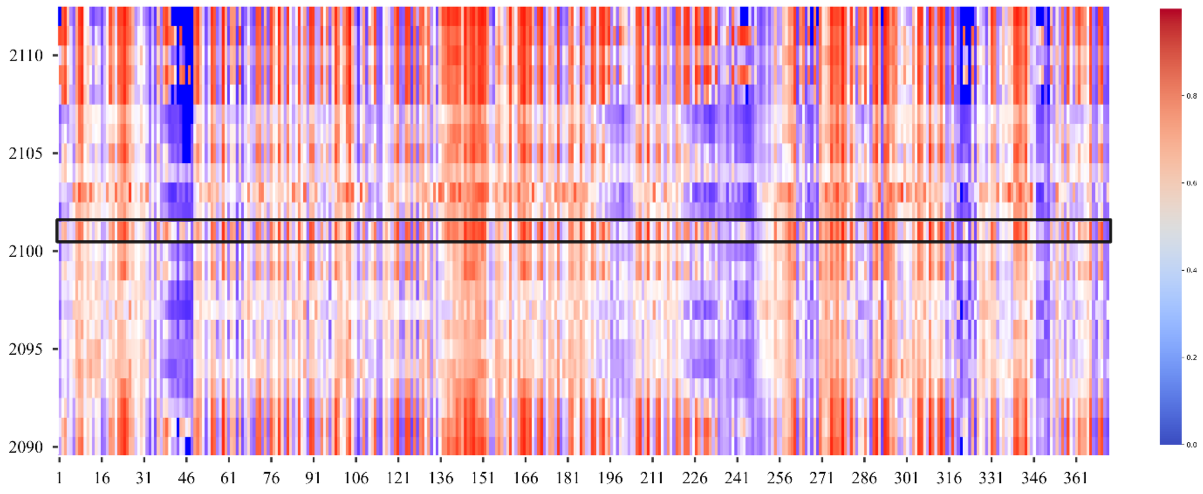

**Figure A20.-** Docknet prediction of *TRIOBP*-5 p.R2101 and *ACTB*

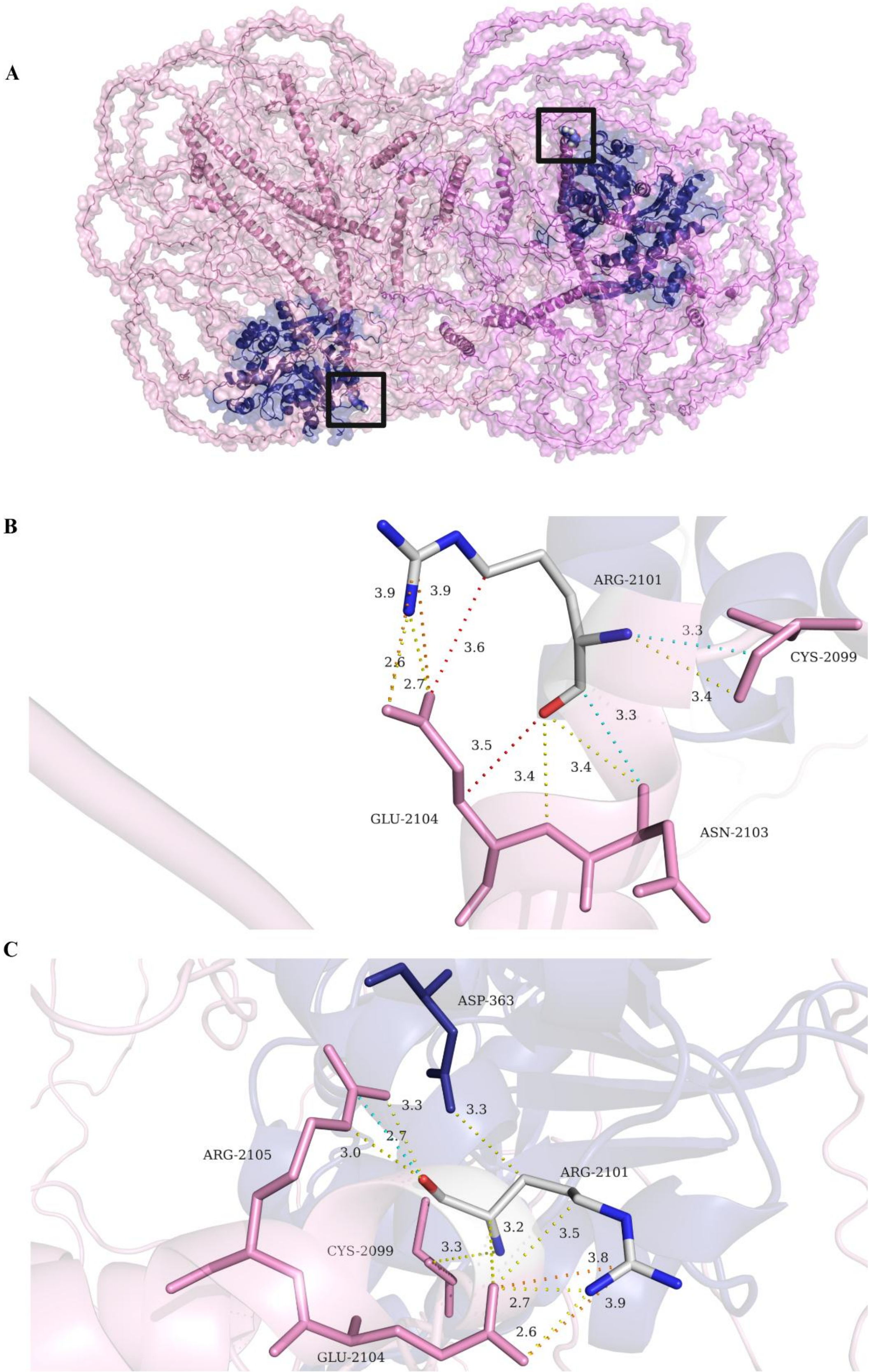

**Figure A21.-** A) Wild type *TRIOBP*-5 dimer docked model to *ACTB* chain C. B) Polar contacts of wt *TRIOBP*-5 dimer docked to *ACTB* chain A. B) Polar contacts of wt *TRIOBP*-5 dimer docked to *ACTB* chain C.

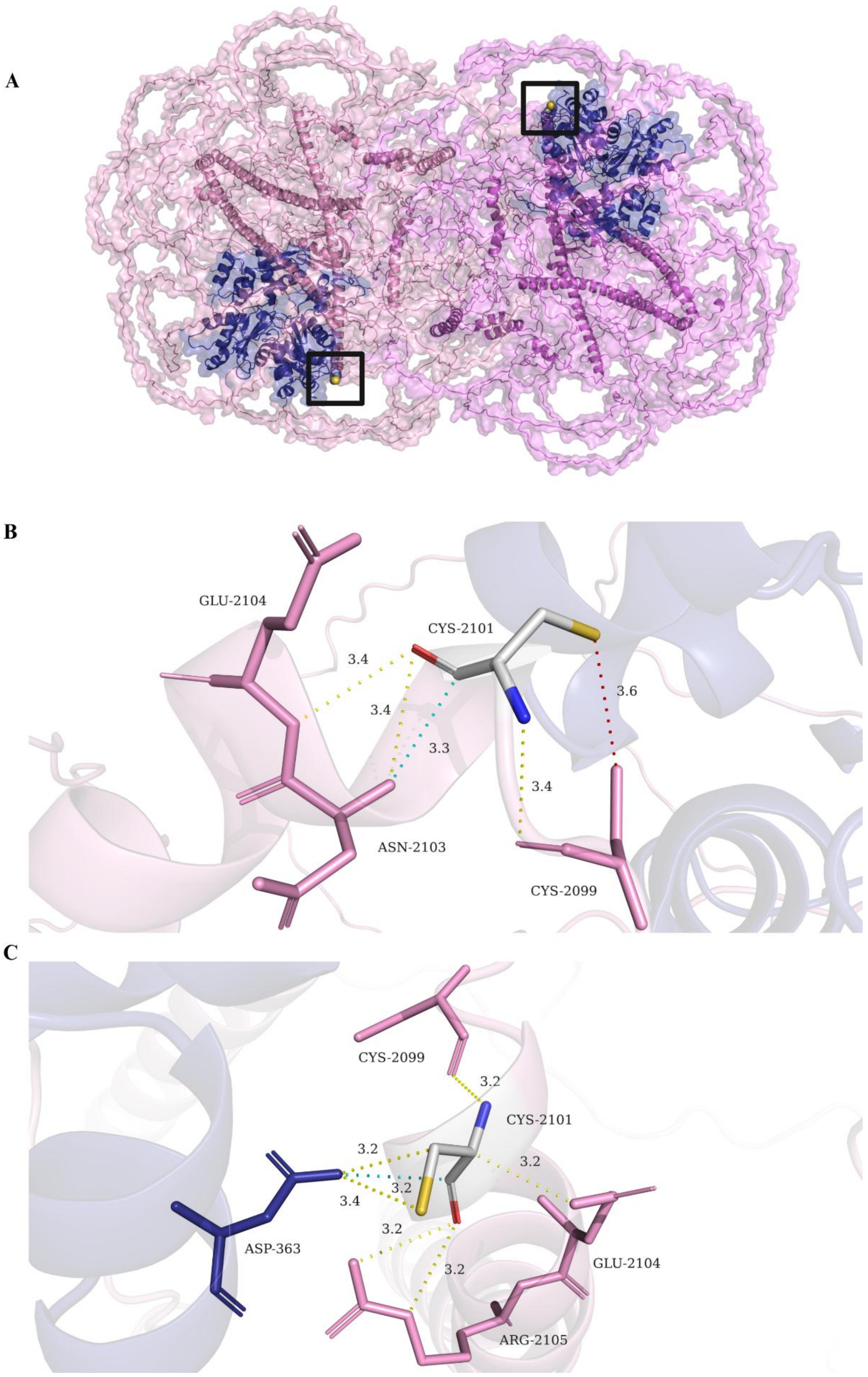

**Figure A22.-** A) Mutant *TRIOBP*-5 dimer docked model to *ACTB* chain C. B) Polar contacts of mutant *TRIOBP*-5 dimer docked to *ACTB* chain A. B) Polar contacts of mutant *TRIOBP*-5 dimer docked to *ACTB* chain C.
